# An economic evaluation of a specialist mental health service for healthcare workers in the East of England

**DOI:** 10.64898/2025.12.19.25342285

**Authors:** Krishnan Puri Sudhir, Rory A Cameron, Adam P Wagner, Sulekha Said, Theodora Karadaki, Cathy Walsh, Peter B Jones, Muzaffer Kaser

**Author notes:** <u>Correspondence</u>: **Muzaffer Kaser**.

## Abstract

**Background:** Healthcare workers experience disproportionately high rates of depression, anxiety, and post-traumatic stress compared with the general population. Within the NHS, work-related stress and mental health–related sickness absence has increased over the past decade, a trend intensified by COVID-19. Mental health support offers are patchy across the UK, and the evidence base around interventions is scarce. The Staff Mental Health Service (SMHS) provides rapid, confidential support for NHS staff across Cambridgeshire and Peterborough. In this study, we report an economic evaluation of this dedicated service.

**Aims:** To assess costs and patient outcomes associated with SMHS treatment, compared with local NHS Talking Therapies (TT) support.

**Method:** A model-based cost–consequence analysis comparing two treatment pathways: SMHS or TT, versus TT only. Routinely collected service data and survey responses informed a decision-tree model estimating costs (2022/23 £GBP), clinical outcomes (PHQ-9 and GAD-7 scores), and quality-adjusted life years (QALYs). Additional analyses examined service waiting times and productivity losses.

**Results:** Costs per patient were slightly higher for SMHS or TT (£614 versus £553), resulting in an incremental cost-effectiveness ratio of £7,126/QALY. Treatment at either SMHS or TT yielded greater improvements in mental health outcomes than TT alone, with mean score reductions of 4.2 versus 2.8 (PHQ-9), and 4.6 versus 2.7 (GAD-7). Median waiting times were substantially shorter at SMHS versus TT from referral to assessment (14 versus 17 days), referral to treatment (22 versus 51 days), and assessment to first treatment (7 versus 30 days; all p<0.001). Productivity losses during waiting periods were lower for SMHS, with an estimated value of £2,018 per patient.

**Conclusions:** The SMHS offers a clinically effective and cost-effective model of support for NHS staff, delivering greater improvements in mental health symptoms, substantially shorter waiting times, and reduced productivity losses at only modest additional cost compared with TT. These findings provide early evidence that specialist services for healthcare workers represent good value for money and support continued investment in specialist staff mental health provision within the NHS.

## Introduction

Healthcare workers are disproportionately affected by mental health conditions such as depression, anxiety, and post-traumatic stress disorder (PTSD) (1). In England, rising rates of burnout and poor well-being amongst National Health Service (NHS) staff have long been identified as a serious and growing concern (2, 3). NHS workers are two to three times more likely than the general working population to require sickness absence due to poor mental health (4). In 2016, the percentage of NHS staff feeling unwell due to work-related stress was 37%, up from 28% in 2008 (5). The COVID-19 pandemic further exacerbated this: a large-scale survey (n=4,378) conducted during the start of the first pandemic wave identified a high prevalence of probable common mental disorders (including depression and anxiety; 58.9%), PTSD (30.2%), and alcohol misuse (10.5%) among healthcare staff, far greater than the prevalence of these disorders observed in the general population (17%, 4.4%, and 1.2% respectively) (6-8).

Demand for better mental health support for the healthcare workforce has grown nationally in response to these challenges. Yet most available support has centred on wellbeing initiatives providing low-intensity support and has not offered the structured assessment or evidence-based treatment needed for mental health conditions requiring clinical intervention. This gap highlights the need for specialist services capable of addressing the substantial unmet mental health needs of healthcare workers, as recognised in several reports (2, 9, 10). One such initiative in the East of England is the Staff Mental Health Service (SMHS), launched by Cambridgeshire and Peterborough NHS Foundation Trust (CPFT) in September 2020 to address growing unmet need in the region. The SMHS provides rapid, free, locally funded, and confidential access to a multidisciplinary team for mental health assessment, treatment, and referral for NHS staff across five trusts (11, 12). A detailed description of the service is provided in Supplementary Materials S1 and S2.

Recent data shows that trends in mental ill health have persisted beyond the pandemic (13). Mental illness has remained the leading cause of sickness absence in the NHS since January 2023, accounting for 23% to 29% of total monthly absences, based on data up to July 2025 (14). The need to address mental health challenges of healthcare workers is more critical than ever (15). Within this context, evaluations are essential to demonstrate not only the clinical effectiveness of such services, but also their cost-effectiveness and sustainability, given financial and capacity constraints facing the NHS. This study conducts an economic evaluation, examining the costs and patient outcomes associated with the SMHS to inform commissioning decisions at regional and national levels.

## Method

### Overview

This study conducted a cost–consequence analysis, incorporating a cost–utility component, using a decision-tree model to compare SMHS treatment with a counterfactual scenario in which staff received care exclusively from the CPFT Talking Therapies (TT) service. While alternative mental health services exist within CPFT (Supplementary Materials S3), expert consultations indicated that TT was the most appropriate comparator given the clinical profile of SMHS patients.

The analysis adopted the perspective of participating NHS provider organisations (Cambridgeshire Community Services NHS Trust, CPFT, Cambridge University Hospitals NHS Foundation Trust, North West Anglia NHS Foundation Trust, and Royal Papworth Hospital NHS Foundation Trust). Accordingly, the analysis included direct, personnel-based costs associated with mental health treatment delivery, specifically staff time and patient contact activities. Medication and other non-personnel costs were excluded owing to inconsistent data availability, with the assumption that these represented a small proportion of total costs. Wider organisational consequences, including waiting times and productivity losses associated with delayed access to treatment were examined separately in supplementary analyses.

The analytic time horizon covered one treatment episode, from first treatment appointment to discharge. Costs incurred before treatment (e.g. triage) or after discharge (e.g. follow-up) were excluded. As treatment duration was generally under one year, discounting was not applied. Results of the cost–utility component, expressed in quality-adjusted life years (QALYs), were reported alongside cost and clinical outcomes.

The study was approved by the NHS Health Research Authority (REC reference: 22/PR/0433, IRAS study ID: 301147) on 11^th^ June 2022 and reported according to the Consolidated Health Economic Evaluation Reporting Standards (CHEERS 2022; Supplementary Material S4) (16).

### Participants and data collection

Two complementary data sources were used: (i) routinely collected, pseudo-anonymised patient-level data from SMHS and CPFT TT, and (ii) pre- and post-treatment questionnaires completed by SMHS patients.

All SMHS patients between November 2022 and April 2024 were informed about the study at triage or assessment and invited to participate. Interested individuals received an information leaflet and provided written consent. Participation was voluntary and withdrawal could occur at any time.

Participants completed identical admission and discharge questionnaires, either online or on paper, capturing sociodemographic and employment details, quality of life measures, and mental health outcomes assessed using the Patient Health Questionnaire-9 (PHQ-9), Generalised Anxiety Disorder-7 scale (GAD-7), and PTSD Checklist–Civilian Version (PCL-C) for depression, anxiety, and PTSD.

Routinely collected data covered all SMHS patients referred between 1 January and 31 December 2022 (given the routine collection, these data were collected irrespective of consent). For TT, data were limited to CPFT patients assessed as suitable for high-intensity treatment, reflecting a clinical profile considered comparable to SMHS users (Supplementary Material S5). Extracted fields included demographics, service contacts from referral to discharge, and PHQ-9, GAD-7, and (for SMHS) PCL-C scores.

After applying exclusion criteria (patients not discharged, unassessed, disengaged, re-referred, aged under 17 or over 65 years, or unemployed at assessment), analytic samples comprised N=2,785 (SMHS: n=446; TT: n=2,339). Of these, n=2,306 (SMHS: n=213; TT: n=2,093) had available resource use data, and n=1,630 (SMHS: n=60; TT: n=1,570) had complete PHQ-9 and GAD-7 data. Flow diagrams presenting identification of patients with available resource use and clinical effectiveness data are provided in Supplementary Materials S6 and S7. Descriptive statistics comparing SMHS subsamples derived from questionnaire versus routinely collected data sources are provided in Supplementary Material S8.

### Analysis

#### Decision-tree model

A decision-tree model was developed in Microsoft Excel to simulate a hypothetical cohort of 1,000 healthcare workers. Model parameters are detailed in Table 1; the model structure is illustrated in Supplementary Material S9. Two treatment alternatives were modelled:

1. **SMHS or TT** – 75% of patients receive SMHS treatment and 25% receive TT.
2. **TT only** – counterfactual wherein all patients are treated exclusively by TT.

**Table 1.** Decision-tree model parameters.

| Parameter | Base case value | Low value | High value | Distribution (parameters) | Reference |
| --- | --- | --- | --- | --- | --- |
| Treatment at either SMHS or Talking Therapies (SMHS or TT) |  |  |  |  |  |
| Probabilities (%) |  |  |  |  |  |
| Patients receiving treatment at SMHS | 75% | 0% | 100% | N/A | Assumption informed by expert input |
| Treatment effectiveness (change in scores) |  |  |  |  |  |
| Depressive symptoms (PHQ-9) |  |  |  |  |  |
| SMHS | -4.7 | -6.1 | -3.3 | Normal ($\sigma=0.7$) | Calculated; see Supplementary Material S10 |
| TT | -2.8 | -3.0 | -2.5 | Normal ($\sigma=0.1$) | |
| Anxiety symptoms (GAD-7) |  |  |  |  |  |
| SMHS | -5.2 | -6.5 | -3.9 | Normal ($\sigma=0.7$) | Calculated; see Supplementary Material S10 |
| TT | -2.7 | -3.0 | -2.5 | Normal ($\sigma=0.1$) | |
| QALY gains |  |  |  |  |  |
| SMHS | 0.34 | 0.33 | 0.35 | Gamma ($\alpha=4786, \beta=0$) | Calculated; see Supplementary Material S11 |
| TT | 0.33 | 0.33 | 0.33 | Gamma ($\alpha=127740, \beta=0$) | |
| Cost of single treatment course (£) |  |  |  |  |  |
| SMHS | £635 | £578 | £692 | Gamma ($\alpha=483, \beta=1.3$) | Calculated; see Supplementary Material S12 |
| TT | £553 | £536 | £571 | Gamma ($\alpha=3954, \beta=0.1$) | |
| Treatment at Talking Therapies only (TT only) |  |  |  |  |  |
| Probabilities (%) |  |  |  |  |  |
| Patients receiving treatment at TT | 100% | N/A |  | N/A | Structural assumption |
| Treatment effectiveness (change in scores) |  |  |  |  |  |
| Depressive symptoms (PHQ-9) |  |  |  |  |  |
| TT | -2.8 | -3.0 | -2.5 | Normal ($\sigma=0.1$) | Calculated; see Supplementary Material S10 |
| Anxiety symptoms (GAD-7) |  |  |  |  |  |
| TT | -2.7 | -3.0 | -2.5 | Normal ($\sigma=0.1$) | Calculated; see Supplementary Material S10 |
| QALY gains |  |  |  |  |  |
| TT | 0.33 | 0.33 | 0.33 | Gamma ($\alpha=127740, \beta=0$) | Calculated; see Supplementary Material S11 |
| Cost of single treatment course (£) |  |  |  |  |  |
| TT | £553 | £536 | £571 | Gamma ($\alpha=3954, \beta=0.1$) | Calculated; see Supplementary Material S12 |

Abbreviations: GAD, Generalised Anxiety Disorder; PHQ, Patient Health Questionnaire; SMHS, Staff Mental Health Service.
Distribution parameters: $\sigma$ , standard error; $\sigma$ , shape; $\beta$ , rate.
N/A indicates that the corresponding parameter does not change in either deterministic (low/high values) or probabilistic (distribution) sensitivity analyses.

Probability inputs for service allocation were informed by SMHS clinician judgement, and uncertainty explored via sensitivity analyses.

Clinical-effectiveness parameters were derived from changes in PHQ-9 and GAD-7 scores between baseline and discharge, adjusted for baseline severity and sociodemographic factors (sex, age, and ethnicity) using ordinary least squares (OLS) regression (Supplementary Material S10).

A cost–utility component estimated changes in health-related quality of life. Utility values were derived by mapping joint PHQ-9 and GAD-7 scores to EQ-5D-3L values using an algorithm developed from national NHS TT service data (17). Individual-level QALY gains were estimated over the treatment period, with adjusted mean QALYs, calculated using OLS regression, incorporated into the model (Supplementary Material S11).

Resource-use data from the routinely collected datasets included staff type, contact mode (face-to-face, telephone, or video), attendance, and duration. Only direct treatment contacts were included; administrative, missed, pre-assessment, and telephone contacts under ten minutes were excluded. Unit costs for staff time were applied using 2021/22 Personal Social Services Research Unit (PSSRU) Unit Costs of Health and Social Care (18). Adjusted mean per-patient costs were estimated using OLS regression controlling for sociodemographic characteristics (sex, age, and ethnicity; Supplementary Material S12). All costs were converted to 2022/23 £GBP values.

Model outputs included mean cost per patient, mean change in PHQ-9 and GAD-7 scores, and mean QALYs gained for each scenario. Incremental differences between SMHS or TT and the counterfactual (TT only) were calculated. An incremental cost-effectiveness ratio (ICER) was also reported as a ratio between incremental costs and incremental QALY gains.

Uncertainty was examined using probabilistic and deterministic sensitivity analyses, and scenario analyses testing key assumptions (use of routinely collected data only; complete-case analysis; and inclusion of only patients meeting caseness thresholds: PHQ-9 ≥10, GAD-7 ≥8, consistent with NHS TT criteria; further detail in Supplementary Material S13).

### Analysis of waiting times and work productivity

Waiting times were compared between SMHS and TT for: referral to assessment; referral to first treatment appointment; and assessment to first treatment appointment. Median times and interquartile ranges were summarised, with between-service differences tested using two-sample Wilcoxon rank-sum tests.

Work productivity was measured using three validated self-report items with a four-week recall period (19, 20). Absenteeism was the number of full workdays missed due to mental health problems; presenteeism was days worked while unwell multiplied by self-rated percentage productivity loss. Total lost productivity equalled the sum of absenteeism and presenteeism-adjusted days, valued using a human-capital approach based on NHS Agenda for Change daily pay (21).

Mean baseline productivity data from the SMHS patients completing the baseline questionnaire were used to estimate losses during treatment waiting times, calculated as ([waiting time / 7] × 5 working days) multiplied by mean daily productivity loss at baseline. It was assumed TT patients would have the same baseline productivity as the SMHS cohort. A conservative scenario assumed an 18% spontaneous remission rate after one month, consistent with the upper range reported in literature (22).

## Results

### Service user characteristics

Service user characteristics are presented in Table 2. These include sociodemographic data and, where available, resource use data, clinical outcomes, and productivity data, reported separately by service.

**Table 2.** Service user characteristics.

|  | Level/statistic | SMHS | Talking Therapies |
| --- | --- | --- | --- |
| Sociodemographic characteristics |  |  |  |
| N |  | 446 | 2,339 |
| Sex | Female | 81% | 71% |
|  | Male | 19% | 29% |
|  | Other | 0% | <1% |
|  | Not known | 0% | <1% |
| Age | Mean | 38 | 37 |
|  | (95% CI) | (37, 39) | (37, 38) |
| Ethnicity | Asian or Asian British | 5% | 4% |
|  | Black or Black British | 2% | 1% |
|  | Mixed | 2% | 3% |
|  | Other | 13% | 1% |
|  | White - British | 44% | 69% |
|  | White - Other | 7% | 10% |
|  | White – Not Specified | 4% | 0% |
|  | Not known | 24% | 12% |
| Resource use data |  |  |  |
| n |  | 213 | 2,093 |
| Therapeutic contacts | Mean number of contacts<br>(95% CI) | 8.9<br>(7.4, 10.4) | 9.5<br>(9.2, 9.7) |
| Clinical outcomes |  |  |  |
| n |  | 60 | 1,570 |
| PHQ-9 (score out of 27) |  |  |  |
| Pre-treatment | Mean score<br>(95% CI) | 17.3<br>(15.7, 18.8) | 12.0<br>(11.7, 12.4) |
| Post-treatment |  | 10.2<br>(8.5, 11.9) | 9.4<br>(9.1, 9.7) |
| GAD-7 (score out of 21) |  |  |  |
| Pre-treatment | Mean score<br>(95% CI) | 14.7<br>(13.5, 15.8) | 11.6<br>(11.3, 11.8) |
| Post-treatment |  | 8.2<br>(6.7, 9.6) | 8.9<br>(8.6, 9.2) |
| Work productivity |  |  |  |
| n |  | 51 |  |
| Work status | Off work | 30% |  |
|  | Not off work | 70% |  |
| Absenteeism<br>(prev. 4 weeks) | Mean days off work<br>(95% CI) | 4.3<br>(2.3, 6.3) |  |
| Presenteeism<br>(prev. 4 weeks) | Mean days working at lower<br>productivity<br>(95% CI) | 10<br>(7.9, 12.1) |  |
|  | Mean self-reported productivity level<sup>1</sup><br>(95% CI) | 51%<br>(45.2%, 56.8%) |  |

All statistics have been rounded to the nearest integer; consequently, percentages may not sum to 100%. Grey boxes indicate data not applicable. <sup>1</sup>Only for participants reporting days worked at lower productivity levels (n=39). Abbreviations: CI, confidence interval; CUH, Cambridge University Hospital; CCS, Cambridgeshire Community Services NHS Trust; CUH, Cambridge University Hospitals NHS Foundation Trust; NWAFT, North West Anglia NHS Foundation Trust; RPH, Royal Papworth Hospital NHS Foundation Trust; SMHS, Staff Mental Health Service.

For both SMHS and TT, similar statistics for sex, age, and ethnicity were observed across services (SMHS: n=446; TT: n=2,339). In both groups, most (∼70-80%) service users were female. Detailed characteristics, including trust of employment and staff type for SMHS patients, are provided in Supplementary Material S14. Among SMHS patients, most were employed at Cambridge University Hospitals (CUH) and CPFT, with approximately one in four working in nursing or midwifery.

Across services, resource use (SMHS: n=213; TT: n=2,093) and clinical effectiveness (SMHS: n=60; TT: n=1,570) characteristics were also comparable. The mean number of treatment appointments did not differ substantially between services, with SMHS patients attending slightly fewer sessions on average (8.9 versus 9.5). At baseline, however, the SMHS sample exhibited higher symptom severity, as indicated by elevated PHQ-9 (17.3 versus 12.0; out of 27) and GAD-7 (14.7 versus 11.6; out of 21) scores compared to the TT sample.

For SMHS patients with complete work productivity data at baseline (n=51), 30% (n=15) reported being off work due to mental illness. The mean number of days taken off due to mental ill health in the previous four weeks was 4.3 (95% CI: 2.3, 6.3). Participants also reported reduced productivity while at work (presenteeism), averaging 10.0 days (95% CI: 7.9, 12.1) over the same period.

### Decision-tree model

#### Base case

Results from the base case analysis are presented in Table 3. For the simulated cohort of 1,000 patients, the total costs per patient for the two treatment options (SMHS or TT and TT only) were £614 and £553, respectively, indicating that SMHS or TT was £61 more expensive per patient compared with TT only.

**Table 3.** Base case and scenario analyses results.

|  | SMHS or<br>Talking Therapies | Talking Therapies<br>only | Difference*/ICER |
| --- | --- | --- | --- |
| **Base case analysis** |  |  |  |
| (Effectiveness: SMHS n = 60; TT n = 1,570 \| Resource use: SMHS n = 213; TT n = 2,093) |  |  |  |
| Costs |  |  |  |
| Cost per patient | £614 | £553 | £61 |
| Effectiveness outputs |  |  |  |
| PHQ-9 | -4.2 | -2.8 | -1.5 |
| GAD-7 | -4.6 | -2.7 | -1.8 |
| QALY gains | 0.34 | 0.33 | 0.01 |
| **QALY ICER** |  |  | **£7,126/QALY gained** |
| **Scenario 1: Routinely collected service-level data only** |  |  |  |
| (Effectiveness: SMHS n = 48; TT n = 1,570 \| Resource use: SMHS n = 201; TT n = 2,093) |  |  |  |
| Cost |  |  |  |
| Cost per patient | £630 | £553 | £76 |
| Effectiveness outputs |  |  |  |
| PHQ-9 | -4.4 | -2.7 | -1.6 |
| GAD-7 | -4.9 | -2.7 | -2.2 |
| QALY gains | 0.34 | 0.33 | 0.01 |
| **QALY ICER** |  |  | **£8,289/QALY gained** |
| **Scenario 2: Complete-case analysis** |  |  |  |
| (SMHS n = 51; TT n = 1,554) |  |  |  |
| Costs |  |  |  |
| Cost per patient | £837 | £547 | £290 |
| Effectiveness outputs |  |  |  |
| PHQ-9 | -4.6 | -2.8 | -1.8 |
| GAD-7 | -5.0 | -2.7 | -2.2 |
| QALY gains | 0.34 | 0.33 | 0.01 |
| **QALY ICER** |  |  | **£28,483/QALY gained** |
| **Scenario 3: Complete-case analysis + ‘caseness’ thresholds for depression and anxiety** |  |  |  |
| (SMHS n = 43; TT n = 896) |  |  |  |
| Costs |  |  |  |
| Cost per patient | £756 | £548 | £208 |
| Effectiveness outputs |  |  |  |
| PHQ-9 | -6.7 | -4.6 | -2.1 |
| GAD-7 | -6.5 | -4.3 | -2.2 |
| QALY gains | 0.32 | 0.32 | <0.01 |
| **QALY ICER** |  |  | **£21,716/QALY gained** |

Costs reported as mean estimates in 2022 £ GBP values.
Outcomes reported as mean score changes.
Base case results include 95% confidence intervals in brackets, estimated through probabilistic sensitivity analysis.
Abbreviations: GAD, Generalised Anxiety Disorder; ICER, Incremental cost-effectiveness ratio; PHQ, Patient Health Questionnaire; QALY, Quality-adjusted life year; SMHS, Staff Mental Health Service; TT, Talking Therapies.

Meanwhile, SMHS or TT resulted in a mean reduction of 4.2 points on the PHQ-9 scale, compared with a reduction of 2.8 points on the same scale for TT only. Similarly, a mean reduction of 4.6 points was observed on the GAD-7 scale for SMHS or TT, versus a reduction of 2.7 points on the same scale for TT only. SMHS or TT, therefore showed an improvement of symptoms compared to TT only on both the PHQ-9 and GAD-7 scales for depression and anxiety, respectively. The resulting (deterministic) base case ICER for SMHS or TT was £7,126 per QALY gained, based on a mean cost difference of £61 and a mean QALY gain of 0.01.

#### Sensitivity analyses

Probabilistic sensitivity analysis from 10,000 iterations of the model indicated that the mean difference in cost per patient between SMHS or TT and TT only was £61 (95% CI: £18, £107). Mean differences in changes in PHQ-9 and GAD-7 scores were -1.5 points (95% CI: -2.5, -0.5) and -1.9 points (95% CI: -2.8, -0.9) respectively. The resulting cost-effectiveness plane and cost-effectiveness acceptability curve are shown in Supplementary Materials S15 and S16, respectively.

Deterministic variation of key parameters revealed that cost associated with SMHS treatment had the largest influence on the difference in cost per patient between SMHS or TT and TT only. Meanwhile, for differences in both PHQ-9 and GAD-7 scores, the proportion of patients treated at SMHS (versus Talking Therapies) in SMHS or TT had the largest influence. Tornado plots are presented in Supplementary Material S17.

#### Scenario analyses

Table 3 reports results from the scenario analyses. Descriptive statistics for patient samples included for calculation of model parameters in each scenario can be found in Supplementary Material S18-S26.

Scenario analyses showed that excluding questionnaire data (Scenario 1) improved incremental score differences compared to the base case (PHQ-9: -1.6 versus -1.5; GAD-7: -2.2 versus -1.8) while also increasing incremental costs (£76 versus £61). Incremental QALY gains remained the same (0.01), corresponding to an increased ICER of £8,289 per QALY gained.

A complete-case analysis (Scenario 2) resulted in a considerably higher incremental cost figure of £290 but also improved incremental score differences (PHQ-9: -1.8; GAD-7: -2.2). Incremental QALY gains remained at 0.01, with the higher incremental costs raising the ICER to £28,483 per QALY gained.

Restricting to patients meeting ‘caseness’ thresholds while maintaining a complete-case analysis (Scenario 3) resulted in an incremental cost of £208 and improved incremental score differences (PHQ-9: -2.1; GAD-7: -2.2). QALY gains again remained at 0.01, with a corresponding ICER of £21,716 per QALY gained.

### Analysis of waiting times and work productivity

#### Waiting times

Waiting times at SMHS were shorter and less variable than at TT. From referral to assessment, the median waiting time at SMHS (n=273) was 14 days (IQR: 10-22; mean: 19), compared to 17 days (IQR: 13-23; mean: 22) at TT (n=2,287). From referral to treatment, waiting times were 22 days (IQR: 15-34; mean: 29) at SMHS (n=273) and 51 days (IQR: 23-95; mean: 66) at TT (n=1,645). Finally, from assessment to the first treatment appointment, waiting times were 7 days (IQR: 5-11; mean: 9) at SMHS (n=273), compared to 30 days (IQR: 2-73; mean: 48) at TT (n=1,508). All differences were statistically significant (p<0.001). Supplementary Material S27 graphically compares waiting times between services using box and whisker plots.

#### Work productivity

Self-reported productivity loss during the waiting period was estimated to average 9.8 working days per patient, equating to an average salary cost of £1,556 (Table 4). Under the counterfactual assumption (staff attended TT, experiencing the same average waiting period, and a constant level of reduced productivity), the base case (no change in mental health status over the waiting period) estimated an average productivity loss of 22.5 days at a salary cost of £3,574, giving a cost difference of £2,018.

**Table 4.** Estimates of mean productivity loss and costs over waiting period (referral to treatment means: SMHS 28.9 days, TT 66.4 days)

| Analysis | Annual salary (£) | Daily rate (£) | Waiting time (working days) |  | Productivity loss (working days) |  | Productivity cost* (salary only, £) |  | Difference |
| --- | --- | --- | --- | --- | --- | --- | --- | --- | --- |
|  |  |  | SMHS | TT | SMHS | TT | SMHS | TT |  |
| Base case | £35,229 | £158.7 | 20.6 | 47.4 | 9.8 | 22.5 | £1,556 | £3,574 | -£2,018 |
| Scenario: 18% remission rate after 1 month |  |  |  |  | 9.8 | 20.3 | £1,556 | £3,221 | -£1,666 |

\*These costs include paid salary only and do not include on-costs (pensions, payroll tax etc.)

Under the more conservative scenario analysis (assuming an 18% spontaneous remission rate after 1 month), productivity loss under TT was reduced to 20.3 working days, and a salary cost of £3,221 (reducing the cost difference between TT and SMHS to £1,666).

Analysis of the relationship between productivity and health status (Supplementary Materials S28-S30) indicated a 2% decrease in productivity with each 1-point increase in both PHQ-9 (p=0.001) and GAD-7 (p=0.06) scores.

## Discussion

This study evaluated the costs and benefits of the Cambridgeshire and Peterborough Staff Mental Health Service (SMHS), a specialist service established to address the mental health needs of NHS staff in the region. The analysis was conducted in the context of other locally available services, particularly NHS Talking Therapies (TT), comparing treatment-related resource use, costs, clinical outcomes, waiting times, and productivity losses. To our knowledge, this is the first economic evaluation of a specialised mental health service for healthcare workers in England.

A decision-tree model was developed to compare the costs and clinical outcomes of two treatment approaches for healthcare workers: access to either SMHS or TT, versus TT alone. While our analysis suggested that SMHS was associated with slightly higher costs, it also appeared to deliver greater improvements in mental health outcomes for both symptoms of anxiety and depression. The base case analysis produced an incremental cost-effectiveness ratio (ICER) of £7,126 per QALY gained, well below the £30,000 per QALY threshold commonly used by the National Institute for Health and Care Excellence (NICE) to identify an alternative as cost-effective (23). These findings were consistent under sensitivity analyses and across a range of scenarios and suggest that the SMHS represents a highly cost-effective intervention.

Relevant comparative literature and research are limited, given few published economic evaluations of specialist mental health services for healthcare workers in England. However, estimates generated for TT in our study are broadly consistent with prior literature. Catarino et al. reported mean per-patient treatment costs for TT ranging from £461 to £685 (2020 values), depending on presenting symptoms (depression or anxiety) and patient severity (24). Similarly, Mukuria et al. estimated mean per-patient costs of £559 (2008 values), alongside average reductions in PHQ-9 and GAD-7 scores of 3.4 and 2.7, respectively, four months after baseline (25). Both studies, however, encompassed a mix of low- and high-intensity TT treatments when assessing costs and, in the case of Mukuria et al., outcomes. In contrast, Radhakrishnan et al. calculated substantially higher average costs of £1,416 per treatment course for high-intensity TT in the East of England (26). This difference likely reflects the use of a top-down costing approach in that study, which aggregated expenditure at the trust level rather than the patient level, thereby capturing a broader range of activities and overheads (e.g. staff training and non–patient-facing work).

The SMHS achieved substantially shorter waiting times than TT across all treatment stages. Delayed access to mental health care is a recognised barrier to recovery and has been associated with poorer clinical outcomes (27). Shorter waiting times at SMHS are therefore likely to contribute meaningfully to the improved clinical outcomes observed. Improved outcomes may also, in part, reflect the fact that the SMHS operates as a multidisciplinary team comprising a psychiatrist, psychologist, mental health nurse, and occupational health nurse. Such a configuration is likely to provide a more comprehensive and coordinated care offer, which may additionally support better clinical outcomes.

Analysis of pre-treatment productivity revealed a considerable burden of lost working time linked to poor mental health among NHS staff. At baseline, participants reported an average loss of nine working days in the previous four weeks, primarily through presenteeism. When modelled across typical waiting periods, the shorter waiting time under the SMHS pathway translated into markedly lower productivity-related cost losses compared with TT, even under conservative assumptions. This modelling assumes that productivity improves following treatment. Although direct validation was not possible in this study, the observed association between productivity and symptom severity, consistent with prior research, supports the plausibility of this assumption (28, 29).

This study has several limitations. First, despite integrating routinely collected and survey data, missingness was substantial, particularly for SMHS outcome measures, limiting sample size and increasing uncertainty.

Second, the comparator population (TT) does not represent a direct equivalent to the SMHS. NHS Talking Therapies operates as a primary care mental health service, whereas SMHS provides specialist secondary service tailored to NHS staff. Although the TT sample was restricted to individuals of working age, in employment, and with similar sociodemographic characteristics; and included only patients receiving high-intensity treatment, differences in baseline symptom severity and unobserved characteristics likely remained.

Third, the analysis required several simplifying assumptions. In the cost–consequence model, certain direct (e.g. triage calls, case discussions) and indirect costs (e.g. training, infrastructure), were excluded, providing a relatively narrow cost perspective. These omissions likely underestimate true service expenditure, although they were applied consistently across both services. Broader organisational consequences of staff sickness, such as administrative, agency, and workload costs, were also not captured.

Lastly, the lack of randomisation and the short analytic time horizon, limited to a single treatment period (which varied between patients), limit causal inference and the assessment of longer-term effects. While these design choices enabled use of real-world data, they may not fully reflect ongoing costs or benefits of treatment, particularly for more severe cases. Future work using larger datasets and dynamic modelling approaches (e.g. Markov or microsimulation models) might address such limitations.

This study provides preliminary evidence that a specialised service for healthcare workers can offer excellent value for money, achieving greater clinical improvement and productivity gains. These findings support continued investment, and further evaluation, in staff mental health services as a cost-effective approach to maintaining workforce stability and service quality in the NHS. Current unmet needs in healthcare workers’ mental health require a multi-level response. A key component of this response is the provision of dedicated mental health services to reduce the treatment gap, which may have downstream benefits for population health by helping maintain the productivity of NHS staff. Our results support the economic case for developing services such as the Staff Mental Health Service in Cambridgeshire and Peterborough.

## Supporting information

SMHS_HE_Supplementary Material

## Acknowledgements

The authors would like to thank the following individuals for their valuable contributions to this study: the service delivery team at CPFT Talking Therapies (James Clarke, Samuel Davies, and Henry Makuyana) for collating and providing service data; Kay Taylor from the Information Governance team at CPFT for support with data protection and sharing; Dawn Brown for assistance in procuring service-level data at SMHS; James Pavey and Charlotte Brigham from the SMHS administration team for their help with data collation; Dr Lorna Bo for support with participant interviews; and David Turner and Prof. Kristy Sanderson from the University of East Anglia for their guidance in refining the health economics and staff productivity methodologies. This research was supported by the NIHR Cambridge Biomedical Research Centre (BRC-1215-20014). The views expressed are those of the author(s) and not necessarily those of the NIHR or the Department of Health and Social Care. Finally, we acknowledge the support of the NIHR Applied Research Collaboration East of England (ARC EoE) throughout the study period.

## Author contribution

RC, PJ, MK, APW, and CW were co-applicants on the original grant application for this research. RC, MK, and APW developed the initial study design. MK, TK, and SS recruited SMHS patients for the study questionnaire. MK and SS collated administrative service-level data from SMHS for analysis. RC, KPS, and APW refined methodology for the economic analysis. KPS built the decision-tree model and conducted associated statistical analyses. RC conducted the work productivity analysis. RC, MK and KPS prepared the initial manuscript with input from APW. APW reviewed the draft and provided additional input. RC and KPS were responsible for drafting the final version of the manuscript, with input from MK and APW. All authors reviewed and approved the final manuscript.

## Transparency declaration

The corresponding author (MK), as manuscript guarantor, affirms that this manuscript is an honest, accurate, and transparent account of the study being reported; that no important aspects of the study have been omitted; and that any discrepancies from the study as planned have been fully explained.

## Data availability

The authors confirm that the data supporting the findings of this study are included within the article. Source data are not publicly available due to the sensitive nature of participant information. Additional details can be requested from the corresponding author (MK).

## Analytic code availability

Analytic code is available from the corresponding author (MK) upon reasonable request.

## Research material availability

Research materials are available from the corresponding author (MK) upon reasonable request.

## References

1. Work-related stress, anxiety or depression statistics in Great Britain, 2024. Health and Safety Executive, 2024.

2. Kinman G, Teoh K, Harriss A. The Mental Health and Wellbeing of Nurses and Midwives in the United Kingdom. The Society of Occupational Medicine (SOM), 2020.

3. Scott HR, Stevelink SAM, Gafoor R, Lamb D, Carr E, Bakolis I, et al. Prevalence of post-traumatic stress disorder and common mental disorders in health-care workers in England during the COVID-19 pandemic: a two-phase cross-sectional study. The Lancet Psychiatry. 2023; 10(1): 40–9.

4. Blaaza M, Shemtob L, Asanati K, Majeed A. A healing challenge: examining NHS staff sickness absence rates. Journal of the Royal Society of Medicine. 2024; 117(2): 55–6.

5. Johnson J, Hall LH, Berzins K, Baker J, Melling K, Thompson C. Mental healthcare staff well-being and burnout: A narrative review of trends, causes, implications, and recommendations for future interventions. International Journal of Mental Health Nursing. 2018; 27(1): 20–32.

6. McManus S, Bebbington PE, Jenkins R, Brugha T. Mental Health and Wellbeing in England: the Adult Psychiatric Morbidity Survey 2014. (eds S McManus, P Bebbington, R Jenkins, T Brugha). NHS Digital, 2016.

7. Lamb D, Gnanapragasam S, Greenberg N, Bhundia R, Carr E, Hotopf M, et al. Psychosocial impact of the COVID-19 pandemic on 4378 UK healthcare workers and ancillary staff: initial baseline data from a cohort study collected during the first wave of the pandemic. Occupational and Environmental Medicine. 2021; 78(11): 801–8.

8. Guidance for Mental Health and Wellbeing Hubs for Health and Social Care Staff. NHS England and Improvement, 2020.

9. Workplace health review. BMJ. 2009.

10. Kinman G, Teoh K. What could make a difference to the mental health of UK doctors? A review of the research evidence.: 2018.

11. Staff Mental Health Service. Cambridgeshire and Peterborough NHS Foundation Trust; 2024. https://www.cpft.nhs.uk/smhs/.

12. Kaser M, Karadaki T, Martin Z, Walsh C. One Year On: Evaluation of the Cambridgeshire and Peterborough Staff Mental Health Service, a Bespoke Mental Health Clinic for Healthcare Workers. BJPsych Open. 2022; 8(S1): S137-S.

13. Work-related ill health and occupational disease in Great Britain. Health and Safety Executive 2025. https://www.hse.gov.uk/statistics/causdis/overview.htm.

14. NHS Sickness Absence Rates. NHS Digital, 2025.

15. Almeida-Meza P, Ledden S, Dempsey B, Smith A, Croak B, Bhundia R, et al. Futureproofing the healthcare workforce in Europe: understanding and addressing psychological distress and occupational outcomes. The Lancet Regional Health – Europe. 2025; 57.

16. Husereau D, Drummond M, Augustovski F, de Bekker-Grob E, Briggs AH, Carswell C, et al. Consolidated Health Economic Evaluation Reporting Standards 2022 (CHEERS 2022) statement: updated reporting guidance for health economic evaluations. BMC Medicine. 2022; 20(1): 23.

17. Franklin M, Hernández Alava M. Enabling QALY estimation in mental health trials and care settings: mapping from the PHQ-9 and GAD-7 to the ReQoL-UI or EQ-5D-5L using mixture models. Quality of Life Research. 2023; 32(10): 2763–78.

18. Unit Costs of Health and Social Care 2022 Manual. Personal Social Services Research Unit, 2023.

19. Sanderson K, Tilse E, Nicholson J, Oldenburg B, Graves N. Which presenteeism measures are more sensitive to depression and anxiety? J Affect Disord. 2007; 101(1): 65–74.

20. Mattke S, Balakrishnan A, Bergamo G, Newberry SJ. A review of methods to measure health-related productivity loss. Am J Manag Care. 2007; 13(4): 211–7.

21. NHS Employers. NHS Terms and Conditions 2023 (Agenda for Change). 2023. https://www.nhsemployers.org/system/files/2023-05/2023-24%20pay%20poster.pdf.

22. Mekonen T, Ford S, Chan GCK, Hides L, Connor JP, Leung J. What is the short-term remission rate for people with untreated depression? A systematic review and meta-analysis. J Affect Disord. 2022; 296: 17–25.

23. NICE health technology evaluations: the manual. (ed NIoHaCE (NICE)): 2022.

24. Catarino A, Harper S, Malcolm R, Stainthorpe A, Warren G, Margoum M, et al. Economic evaluation of 27,540 patients with mood and anxiety disorders and the importance of waiting time and clinical effectiveness in mental healthcare. Nature Mental Health. 2023; 1(9): 667–78.

25. Mukuria C, Brazier J, Barkham M, Connell J, Hardy G, Hutten R, et al. Cost-effectiveness of an Improving Access to Psychological Therapies service. The British Journal of Psychiatry. 2013; 202(3): 220–7.

26. Radhakrishnan M, Hammond G, Jones PB, Watson A, McMillan-Shields F, Lafortune L. Cost of Improving Access to Psychological Therapies (IAPT) programme: An analysis of cost of session, treatment and recovery in selected Primary Care Trusts in the East of England region. Behaviour Research and Therapy. 2013; 51(1): 37–45.

27. van Dijk DA, Meijer RM, van den Boogaard TM, Spijker J, Ruhé HG, Peeters FPML. Worse off by waiting for treatment? The impact of waiting time on clinical course and treatment outcome for depression in routine care. J Affect Disord. 2023; 322: 205–11.

28. Beck A, Crain LA, Solberg LI, Unützer J, Maciosek MV, Whitebird RR, et al. The effect of depression treatment on work productivity. Am J Manag Care. 2014; 20(8): e294–301.

29. de Oliveira C, Saka M, Bone L, Jacobs R. The Role of Mental Health on Workplace Productivity: A Critical Review of the Literature. Appl Health Econ Health Policy. 2023; 21(2): 167–93.

