## Supplementary material for "An economic evaluation of a specialist mental health service for healthcare workers in the East of England": SMHS_HE_Supplementary Material

#### S1. SMHS service description.

The Staff Mental Health Service (SMHS) was established in September 2020 to provide rapid access to mental health assessment, treatment and referral for NHS staff working in the five NHS trusts across Cambridgeshire and Peterborough (1):

- Cambridgeshire and Peterborough NHS Foundation Trust (CPFT);
- Cambridgeshire Community Services NHS Trust (CCS);
- Cambridge University Hospitals NHS Foundation Trust (CUH);
- North West Anglia NHS Foundation Trust (NWAFT);
- Royal Papworth Hospital NHS Foundation Trust (RPH).

The service provides free and confidential access to a multidisciplinary team of trained Mental Health Nurses, Occupational Health Specialists, Clinical Psychologists, and Consultant Psychiatrists. For the initial referral to the service, staff must be referred by one of the following (1):

- Their general practitioner (GP);
- Occupational health service at their employing trust;
- Staff support and wellbeing services;
- Any other mental health service within CPFT, such as Primary Care Mental Health (PCMH), Talking Therapies, or Adult Locality Teams (ALTs).

Once the referral is received, the SMHS team endeavours to make first contact with the patient within 72 hours. During this contact, the patient is triaged, and those deemed more suitable for other mental health services are referred or signposted elsewhere. If the patient is assessed eligible for the service, they are offered an ‘initial assessment’, during which a Consultant Psychiatrist or a Clinical Nurse Specialist further investigates their needs and discusses a tailored treatment plan with them. Treatment plans are highly individualised and tailored to the patient’s specific needs; they may involve any combination of:

- Psychiatric input and review provided by a Consultant Psychiatrist;
- Brief psychological interventions provided by specialist Mental Health Nurses, usually over 8 to 12 sessions or 3 to 4 months;
- Talking therapy provided by a Clinical Psychologist, usually over 20 sessions or 6 months.

Once a treatment course is complete, the team assesses whether to continue treatment or discharge the patient. Should further needs arise, anyone who had input from SMHS can self-refer within 6 months; a re-referral from a professional is needed beyond 6 months.

#### S2. SMHS patient pathway.

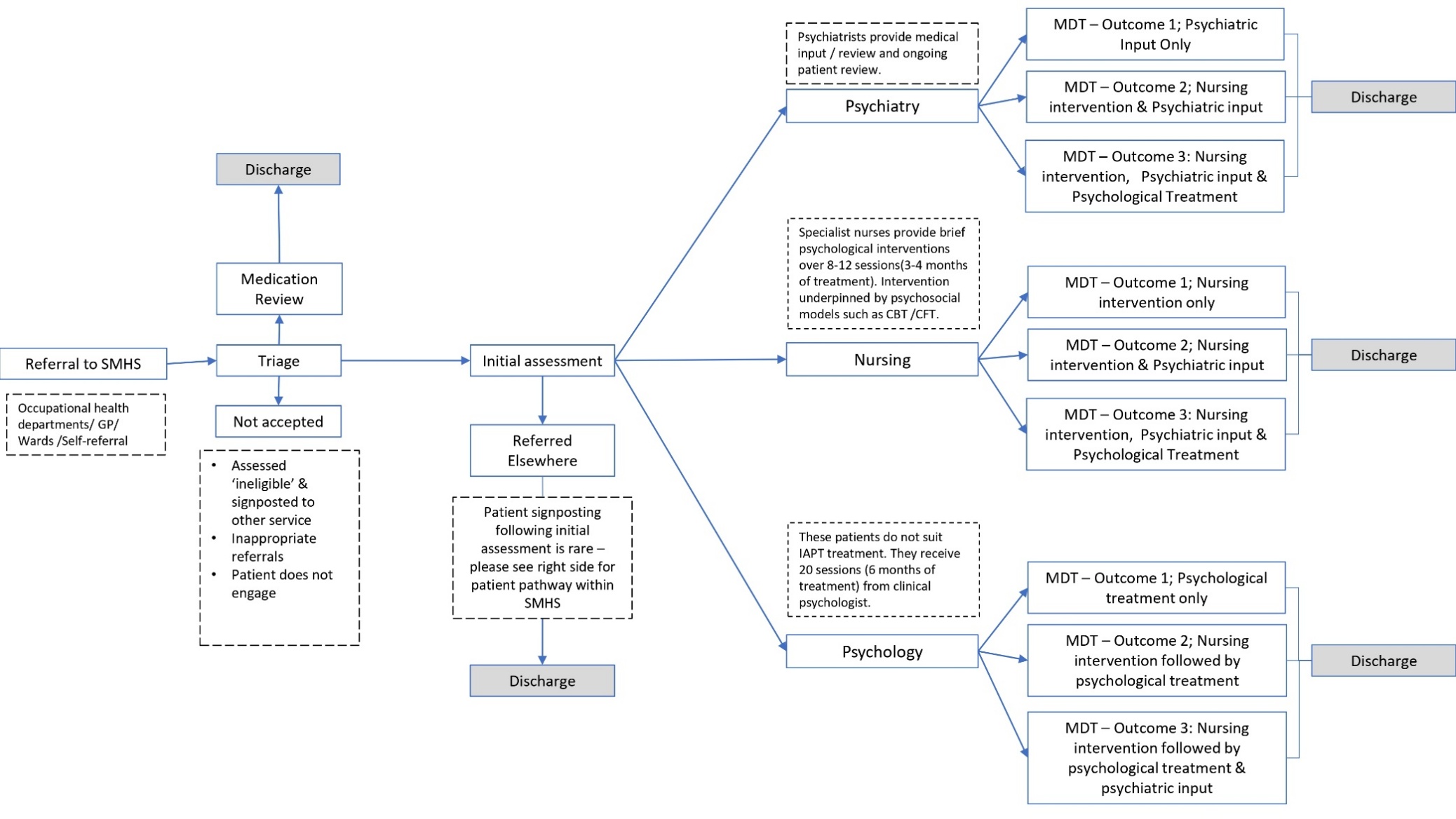

Abbreviations: CBT, Cognitive behavioural therapy; CFT, Compassion focused therapy; GP, general practitioner; IAPT, Improving Access to Psychological Therapies (now called ‘NHS Talking Therapies’); MDT, Multidisciplinary team; SMHS, Staff Mental Health Service.

#### S3. Overview of CPFT mental health care provision.

The main provider of mental health services in Cambridgeshire and Peterborough is the CPFT (2). As elsewhere in England, mental health services are structured into three tiers. Primary care mental health services are provided through two major routes:

- General practitioners, supported in practice by a Primary Care Mental Health (PCMH) team;
- The nationally organised NHS ‘Talking Therapies’ programme, NHS England’s flagship initiative (formerly referred to as Improving Access to Psychological Therapies, IAPT) to deliver psychological therapy to patients aged 17 and over with depression and anxiety disorders.

Once in contact with primary care services, patients may be referred to secondary mental health care services for assessment and treatment if cases are too severe or complex to be appropriately addressed within primary care alone. These services comprise a range of community-based teams, including the SMHS, established to cater to different needs. Examples of local services within this tier include:

- Adult Locality Teams (ALTs), a set of four interdisciplinary teams throughout the area (located in Huntingdon, Fenland, Peterborough and Borders, and Cambridge) providing health and social care to people aged 16 and over, with moderate to severe mental health problems;
- CAMEO (Cambridgeshire and Peterborough Assessing, Managing and Enhancing Outcomes), a service for patients aged between 14 and 35 with psychosis symptoms;
- Inpatient hospital services for adults, including mental health wards (Mulberry, Oak, and Poplar wards);
- Other services, such as the First Response Service (FRS), the Crisis Resolution and Home Treatment Team (CRHTT), and the Individual Placement Support (IPS) service.

The third tier consists of tertiary mental health services: highly specialised services typically set up regionally or nationally to address specific patient populations. Examples of these available in C&P include:

- The Community Forensic Team, a county-wide service that provides mental healthcare to patients who pose a significant risk of causing harm to others.
- The Fens Unit, a team at HMP Whitemoor that addresses the mental health needs of prisoners with severe personality disorders.
- Springbank, an inpatient recovery unit for women suffering from borderline personality disorder (BPD).

Secondary and tertiary services listed above are not exhaustive, and a full list of adult mental health services offered in Cambridgeshire and Peterborough can be viewed on the CPFT website (3).

#### S4. CHEERS 2022 checklist (4).

| **Topic** | **No.** | **Item** | **Location where item is reported** |
| --- | --- | --- | --- |
| **Title** | 1 | Identify the study as an economic evaluation and specify the interventions being compared. | Title |
| **Abstract** | 2 | Provide a structured summary that highlights context, key methods, results, and alternative analyses. | Abstract (structured) |
| **Introduction** | | | |
| **Background and objectives** | 3 | Give the context for the study, the study question, and its practical relevance for decision making in policy or practice. | Introduction section |
| **Methods** | | | |
| **Health economic analysis plan** | 4 | Indicate whether a health economic analysis plan was developed and where available. | A health economics plan was not developed. |
| **Study population** | 5 | Describe characteristics of the study population (such as age range, demographics, socioeconomic, or clinical characteristics). | Method section, under ‘Participants and data collection’; Results section, under ‘Service user characteristics’ |
| **Setting and location** | 6 | Provide relevant contextual information that may influence findings. | Introduction section; Method section, under ‘Overview’ |
| **Comparators** | 7 | Describe the interventions or strategies being compared and why chosen. | Method section, under ‘Overview’ |
| **Perspective** | 8 | State the perspective(s) adopted by the study and why chosen. | Method section, under ‘Overview’ |
| **Time horizon** | 9 | State the time horizon for the study and why appropriate. | Method section, under ‘Overview’ |
| **Discount rate** | 10 | Report the discount rate(s) and reason chosen. | Method section, under ‘Overview’ |
| **Selection of outcomes** | 11 | Describe what outcomes were used as the measure(s) of benefit(s) and harm(s). | Method section, under ‘Analysis’ |
| **Measurement of outcomes** | 12 | Describe how outcomes used to capture benefit(s) and harm(s) were measured. | Method section, under ‘Analysis’ |
| **Valuation of outcomes** | 13 | Describe the population and methods used to measure and value outcomes. | Method section, under ‘Analysis’ (QALY mapping to EQ-5D-3L) |
| **Measurement and valuation of resources and costs** | 14 | Describe how costs were valued. | Method section, under ‘Analysis’ (PSSRU unit costs) |
| **Currency, price date, and conversion** | 15 | Report the dates of the estimated resource quantities and unit costs, plus the currency and year of conversion. | Method section, under ‘Analysis’ |
| **Rationale and description of model** | 16 | If modelling is used, describe in detail and why used. Report if the model is publicly available and where it can be accessed. | Method section, under ‘Overview’ and ‘Analysis, Decision tree model’; Supplementary Materials S9 |
| **Analytics and assumptions** | 17 | Describe any methods for analysing or statistically transforming data, any extrapolation methods, and approaches for validating any model used. | Method section, under ‘Analysis’; Supplementary Materials S9-11 |
| **Characterising heterogeneity** | 18 | Describe any methods used for estimating how the results of the study vary for subgroups. | Method section, under ‘Analysis’; Supplementary Material S12 |
| **Characterising distributional effects** | 19 | Describe how impacts are distributed across different individuals or adjustments made to reflect priority populations. | N/A, not explored |
| **Characterising uncertainty** | 20 | Describe methods to characterise any sources of uncertainty in the analysis. | Method section, under ‘Analysis’ |
| **Approach to engagement with patients and others affected by the study** | 21 | Describe any approaches to engage patients or service recipients, the general public, communities, or stakeholders (such as clinicians or payers) in the design of the study. | N/A |
| **Results** | | | |
| **Study parameters** | 22 | Report all analytic inputs (such as values, ranges, references) including uncertainty or distributional assumptions. | Table 1 |
| **Summary of main results** | 23 | Report the mean values for the main categories of costs and outcomes of interest and summarise them in the most appropriate overall measure. | Results section |
| **Effect of uncertainty** | 24 | Describe how uncertainty about analytic judgments, inputs, or projections affect findings. Report the effect of choice of discount rate and time horizon, if applicable. | Results section, under ‘Decision-tree model’, subsection ‘Sensitivity analyses’; Results section, under ‘Decision-tree model’, subsection Scenario analyses; Supplementary Materials S15-S17 |
| **Effect of engagement with patients and others affected by the study** | 25 | Report on any difference patient/service recipient, general public, community, or stakeholder involvement made to the approach or findings of the study | N/A |
| **Discussion** | | | |
| **Study findings, limitations, generalisability, and current knowledge** | 26 | Report key findings, limitations, ethical or equity considerations not captured, and how these could affect patients, policy, or practice. | Discussion |
| **Other relevant information** | | | |
| **Source of funding** | 27 | Describe how the study was funded and any role of the funder in the identification, design, conduct, and reporting of the analysis | ‘Funding’ statement at end of manuscript |
| **Conflicts of interest** | 28 | Report authors conflicts of interest according to journal or International Committee of Medical Journal Editors requirements. | ‘Declaration of interest’ statement at end of manuscript |

#### S5. **Clinical profile of SMHS and TT samples with available GAD-7 and PHQ-9 data, before and after treatment.**

| **PHQ-9 score severity** | **SMHS (% of patients)** | | **Talking Therapies (% of patients)** | |
| --- | --- | --- | --- | --- |
|  | *Pre-treatment* | *Post-treatment* | *Pre-treatment* | *Post-treatment* |
| **n** | 60 | | 1,570 | |
| **Minimal or no symptoms**  (0-4) | 3% | 25% | 13% | 25% |
| **Mild**  (5-9) | 10% | 30% | 24% | 30% |
| **Moderate**  (10-14) | 18% | 15% | 29% | 25% |
| **Moderately severe**  (15-19) | 28% | 17% | 20% | 12% |
| **Severe**  (20-27) | 40% | 13% | 14% | 8% |

| **GAD-7 score severity** | **SMHS (% of patients)** | | **Talking Therapies (% of patients)** | |
| --- | --- | --- | --- | --- |
|  | *Pre-treatment* | *Post-treatment* | *Pre-treatment* | *Post-treatment* |
| **n** | 60 | | 1,570 | |
| **Minimal or no symptoms**  (0-4) | 3% | 32% | 12% | 26% |
| **Mild**  (5-9) | 13% | 33% | 27% | 32% |
| **Moderate**  (10-14) | 30% | 13% | 27% | 23% |
| **Severe**  (15-21) | 53% | 22% | 35% | 19% |

Abbreviations: GAD; Generalised Anxiety Disorder; PHQ, Patient Health Questionnaire; SMHS, Staff Mental Health Service.

All statistics have been rounded to the nearest integer; consequently, percentages may not sum to 100%.

#### S6. Flow diagram showing patient sample identification for calculation of resource use data.

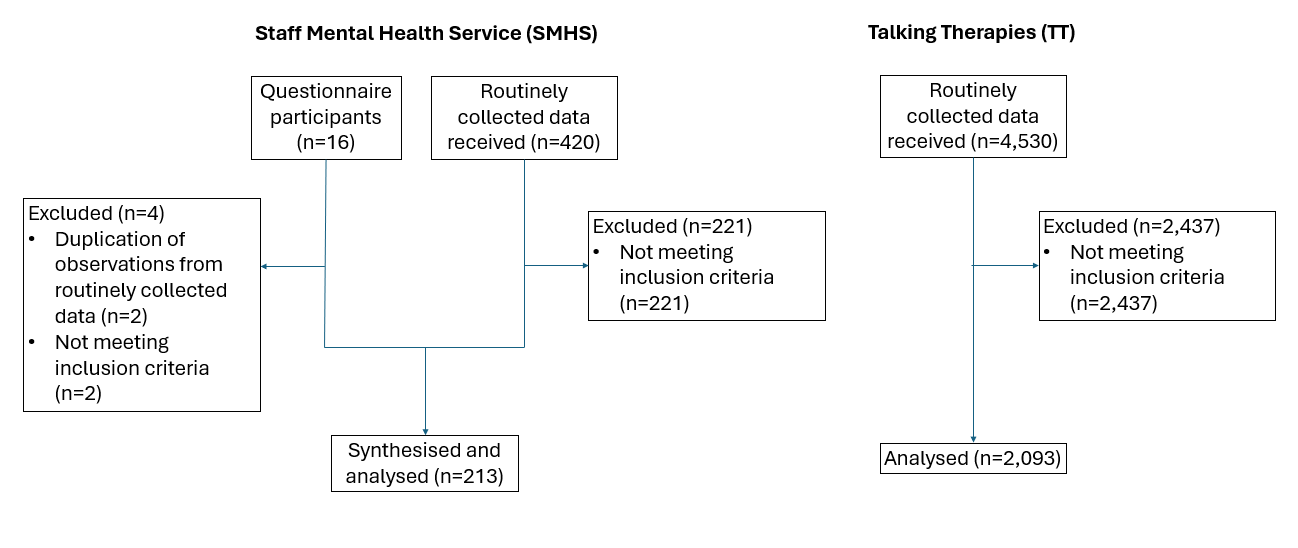

#### S7. Flow diagram showing patient sample identification for calculation of clinical effectiveness (PHQ-9, GAD-7) and utility data.

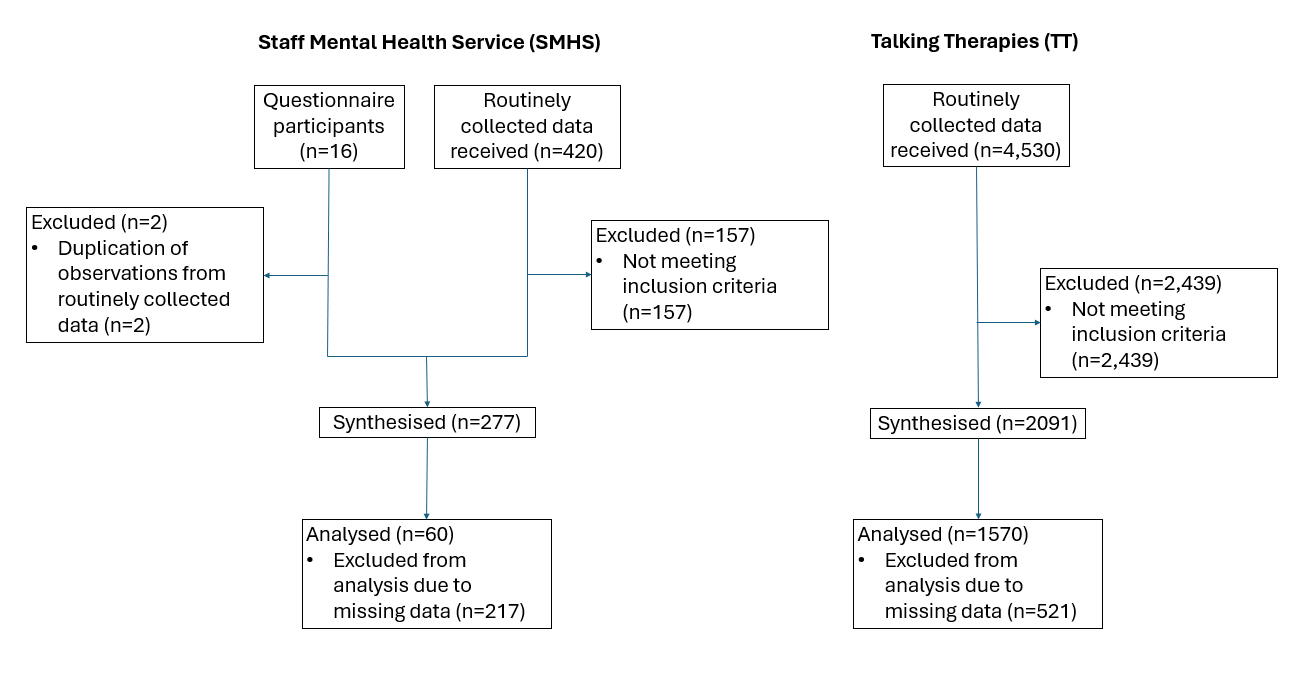

#### S8. Descriptive statistics comparing SMHS subsamples derived from questionnaire versus routinely collected data

|  | **Level/statistic** | **Questionnaire data** | **Routinely collected data** |
| --- | --- | --- | --- |
| **Sociodemographic characteristics** | | | |
| N |  | 51 | 395 |
| Sex | Female | 82.35% | 80.76% |
|  | Male | 17.65% | 19.24% |
| Age | Mean  (95% CI) | 36.73 (33.70, 39.75) | 37.93 (36.76, 39.11) |
| Ethnicity | Asian or Asian British | 1.96% | 5.82% |
|  | Black or Black British | 0.00% | 2.03% |
|  | Mixed | 0.00% | 2.03% |
|  | Other | 0.00% | 14.68% |
|  | White - British | 11.76% | 47.85% |
|  | White - Other | 17.65% | 5.57% |
|  | White – Not Specified | 33.33% | 0.00% |
|  | Not known | 35.29% | 22.03% |
| **Resource use data** | | | |
| n |  | 12 | 201 |
| Therapeutic contacts | Mean number of contacts  (95% CI) | 3.42 (0.98, 5.85) | 9.23  (7.65, 10.81) |
| **Clinical outcomes** | | | |
| n |  | 12 | 48 |
| *PHQ-9 (score out of 27)* | | | |
| Pre-treatment | Mean score (95% CI) | 18.75 (16.40, 21.10) | 16.90 (15.05, 18.74) |
| Post-treatment |  | 11.83 (7.33, 16.34) | 9.81  (7.91, 11.72) |
| *GAD-7 (score out of 21)* | | | |
| Pre-treatment | Mean score (95% CI) | 16.58 (14.04, 19.13) | 14.19 (12.85, 15.53) |
| Post-treatment |  | 10.83 (6.56, 15.10) | 7.50 (6.00, 9.00) |

Grey boxes indicate data not applicable.

Sex variable is dichotomised as ‘Female’ or ‘Male’ by NHS data systems.

Abbreviations: CI, confidence interval.

#### S9. Decision-tree model structure

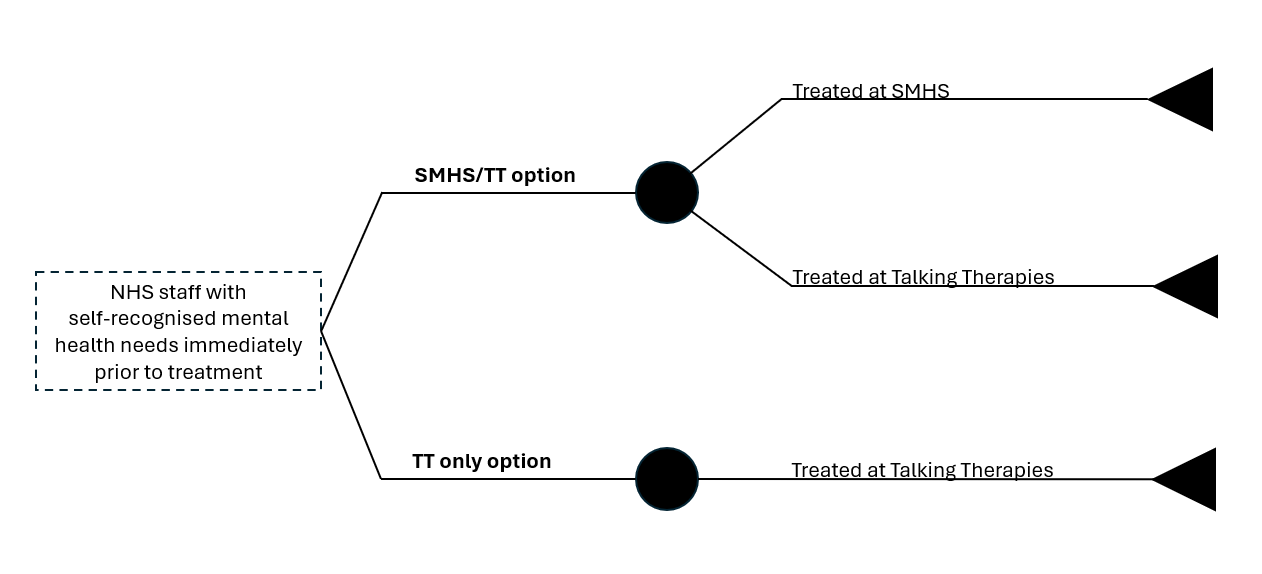

Abbreviations: NHS, National Health Service; SMHS, Staff Mental Health Service; TT, Talking Therapies.

#### S10. Regression models used to estimate treatment effects.

| **Variable** | **OLS model #1: PHQ-9 score** | | **OLS model #2: GAD-7 score** | |
| --- | --- | --- | --- | --- |
|  | **Coefficient** | **p-value** | **Coefficient** | **p-value** |
| TT group (SMHS as reference) | -1.94*** | 0.007 | -2.46*** | <0.001 |
| Score at baseline (pre-treatment) | 0.46*** | <0.001 | 0.47*** | <0.001 |
| Age | 0.02 | 0.200 | 0.00 | 0.998 |
| Sex  (Female as reference) |  |  |  |  |
| *Not known* | 1.00 | 0.848 | -9.69 | 0.046 |
| *Male* | 0.14 | 0.632 | 0.30 | 0.163 |
| Ethnicity (Asian or Asian British as reference) |  |  |  |  |
| *Black or Black British* | -2.24 | 0.112 | -1.86 | 0.156 |
| *Mixed* | -1.33 | 0.184 | -1.42 | 0.125 |
| *White - British* | -0.53 | 0.450 | -0.84 | 0.192 |
| *White - Other* | -0.89 | 0.262 | -1.00 | 0.171 |
| *Other* | -0.42 | 0.735 | -0.30 | 0.791 |
| *Not known* | -0.35 | 0.669 | -1.37 | 0.073 |
| Constant | -1.00 | | 0.53 | |
| Observations (n) | 1,630 | | | |
| R-squared | 0.25 | | 0.23 | |

Abbreviations: GAD; Generalised Anxiety Disorder; OLS, ordinary least squares; PHQ, Patient Health Questionnaire; TT, Talking Therapy.

*** p<0.01, ** p<0.05, * p<0.1

#### S11. Regression model used to estimate QALY differences.

| **Variable** | **OLS model: QALY gains** | |
| --- | --- | --- |
|  | **Coefficient** | **p-value** |
| TT group (SMHS as reference) | -0.01** | 0.024 |
| Treatment duration | 0.68*** | <0.001 |
| Utility at baseline (pre-treatment) | 0.35*** | <0.001 |
| Age | <-0.01*** | 0.005 |
| Sex  (Female as reference) |  |  |
| *Not known* | -0.10 | 0.010 |
| *Male* | <0.01*** | 0.004 |
| Ethnicity (Asian or Asian British as reference) |  |  |
| *Black or Black British* | -0.01 | 0.581 |
| *Mixed* | -0.01 | 0.331 |
| *White - British* | -0.01 | 0.173 |
| *White - Other* | -0.01 | 0.074 |
| *Other* | -0.01 | 0.298 |
| *Not known* | -0.01* | 0.070 |
| Constant | -0.20 | |
| Observations (n) | 1,630 | |
| R-squared | 0.96 | |

Abbreviations: OLS, ordinary least squares; TT, Talking Therapy; QALY, quality-adjusted life year.

*** p<0.01, ** p<0.05, * p<0.1

#### **S12.** Regression model used to estimate treatment cost differences.

| **Variable** | **OLS model: Cost per patient** | |
| --- | --- | --- |
|  | **Coefficient** | **p-value** |
| TT group (SMHS as reference) | -81.44*** | 0.008 |
| Age | 0.86 | 0.270 |
| Sex  (Female as reference) |  |  |
| *Not known* | 129.22 | 0.648 |
| *Male* | -49.90*** | 0.009 |
| Ethnicity (Asian or Asian British as reference) |  |  |
| *Black or Black British* | -135.73 | 0.113 |
| *Mixed* | 115.46* | 0.073 |
| *White - British* | -13.53 | 0.742 |
| *White - Other* | 37.64 | 0.434 |
| *Other* | -28.90 | 0.683 |
| *Not known* | -51.23 | 0.267 |
| Constant | 627.30 | |
| Observations (n) | 2,306 | |
| R-squared | 0.00 | |

Abbreviations: OLS, ordinary least squares; TT, Talking Therapy.

*** p<0.01, ** p<0.05, * p<0.1

#### S13. Detailed overview of scenario analyses

A range of scenarios were developed to explore model assumptions and uncertainty in utilised data:

- **Scenario #1 – routinely collected service-level data only:** To consider any heterogeneity between these two samples, this scenario explored the impact of only analysing service users with routinely collected data^[1]^. In the base case analysis, both questionnaire and routinely collected service-level data for the SMHS were synthesised into one sample to calculate treatment effectiveness and resource use.
- **Scenario #2 – complete-case analysis:** This scenario explored the impact of restricting the analysis to service users with both treatment effectiveness and resource use data (i.e., a complete-case analysis). In the base case, different samples of patients were used to calculate treatment effectiveness and resource use, respectively. For both services, treatment effectiveness was only calculated across patients with reported baseline and follow-up PHQ-9 and GAD-7 scores, regardless of missing resource use data. Similarly, resource use was calculated across patients regardless of missing outcome data.
- **Scenario #3 – complete-case analysis + ‘caseness’ thresholds for symptoms of depression and anxiety:** This scenario was undertaken to reflect clinical decision-making at TT and improve comparability to published TT standards. Thresholds of 10 on the PHQ-9 scale and 8 on the GAD-7 scale were used to indicate ‘caseness’, as employed by Talking Therapies (5). ‘Caseness’ is a term used within clinical practice at NHS Talking Therapies for when a referral is assessed as being a ‘clinical case’. In the base case, diagnostic cut-offs for symptom severity on the PHQ-9 and GAD-7 scales were not incorporated in sample selection for the calculation of clinical effectiveness and resource use parameters in the model. A scenario was considered where in addition to a complete-case analysis, only patients meeting thresholds for diagnostic ‘caseness’ for depression and anxiety (and above) on the PHQ-9 and GAD-7 measures, respectively, were analysed for both clinical effectiveness and resource use.

#### S14. Detailed sociodemographic characteristics for final analytic sample.

|  | **Level/statistic** | **SMHS** | **Talking Therapies** |
| --- | --- | --- | --- |
| **N** |  | 446 | 2,339 |
| **Sex** | Female | 80.94% | 70.97% |
|  | Male | 19.06% | 28.52% |
|  | Other | 0.00% | 0.30% |
|  | Not known | 0.00% | 0.21% |
| **Age** | Mean  (95% CI) | 37.79 (36.70, 38.89) | 37.32 (36.88, 37.76) |
| **Ethnicity** | Asian or Asian British | 5.38% | 4.23% |
|  | Black or Black British | 1.79% | 1.07% |
|  | Mixed | 1.79% | 2.99% |
|  | Other | 13.00% | 12.31% |
|  | White - British | 43.72% | 68.79% |
|  | White - Other | 6.95% | 9.75% |
|  | White – Not Specified | 3.81% | 0.00% |
|  | Not known | 23.54% | 0.86% |
| **NHS trust** | CUH | 46.86% |  |
|  | CCS | 1.79% |  |
|  | CPFT | 27.13% |  |
|  | NWAFT | 16.60% |  |
|  | RPH | 6.73% |  |
|  | Other | 0.45% |  |
|  | Not known | 0.45% |  |
| **Staff group** | Additional Clinical Services | 18.69% |  |
|  | Additional Professional Scientific and Technical | 5.86% |  |
|  | Admin and Clerical | 14.41% |  |
|  | Allied Health Professional | 13.96% |  |
|  | Estates and Ancillary | 2.48% |  |
|  | Healthcare Scientists | 0.90% |  |
|  | Medical and Dental | 7.21% |  |
|  | Nursing and Midwifery | 23.87% |  |
|  | Not known | 12.62% |  |

Grey boxes indicate data not applicable.

Sex variable is dichotomised as ‘Female’ or ‘Male’ by NHS data systems.

Abbreviations: CI, confidence interval; CUH, Cambridge University Hospital; CCS, Cambridgeshire Community Services NHS Trust; CUH, Cambridge University Hospitals NHS Foundation Trust; NWAFT, North West Anglia NHS Foundation Trust; RPH, Royal Papworth Hospital NHS Foundation Trust; SMHS, Staff Mental Health Service.

#### S15. Cost-effectiveness plane (probabilistic sensitivity analysis).

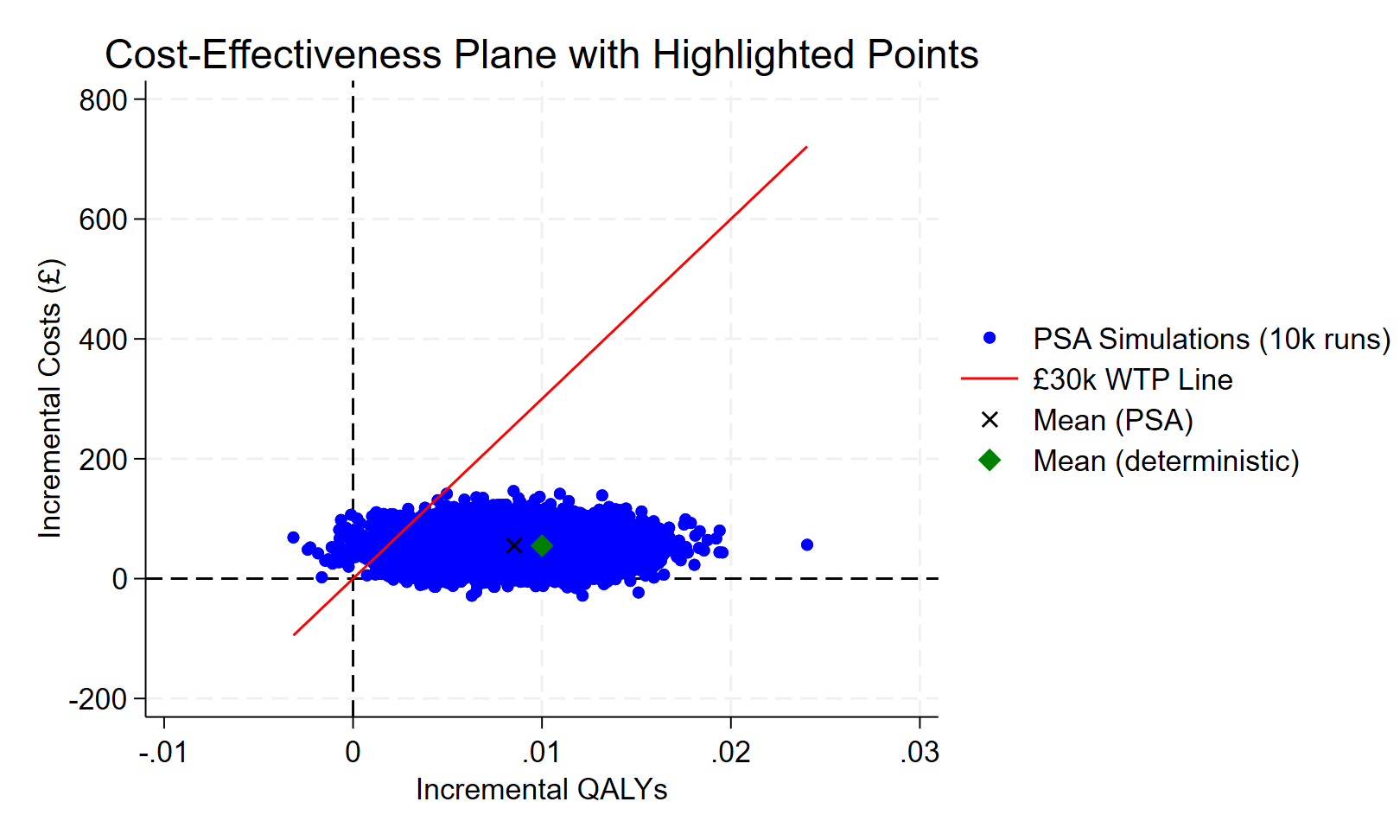

Abbreviations: PSA, probabilistic sensitivity analysis; QALYs, quality-adjusted life years; WTP, willingness-to-pay.

#### S16. Cost-effectiveness acceptability curve (probabilistic sensitivity analysis).

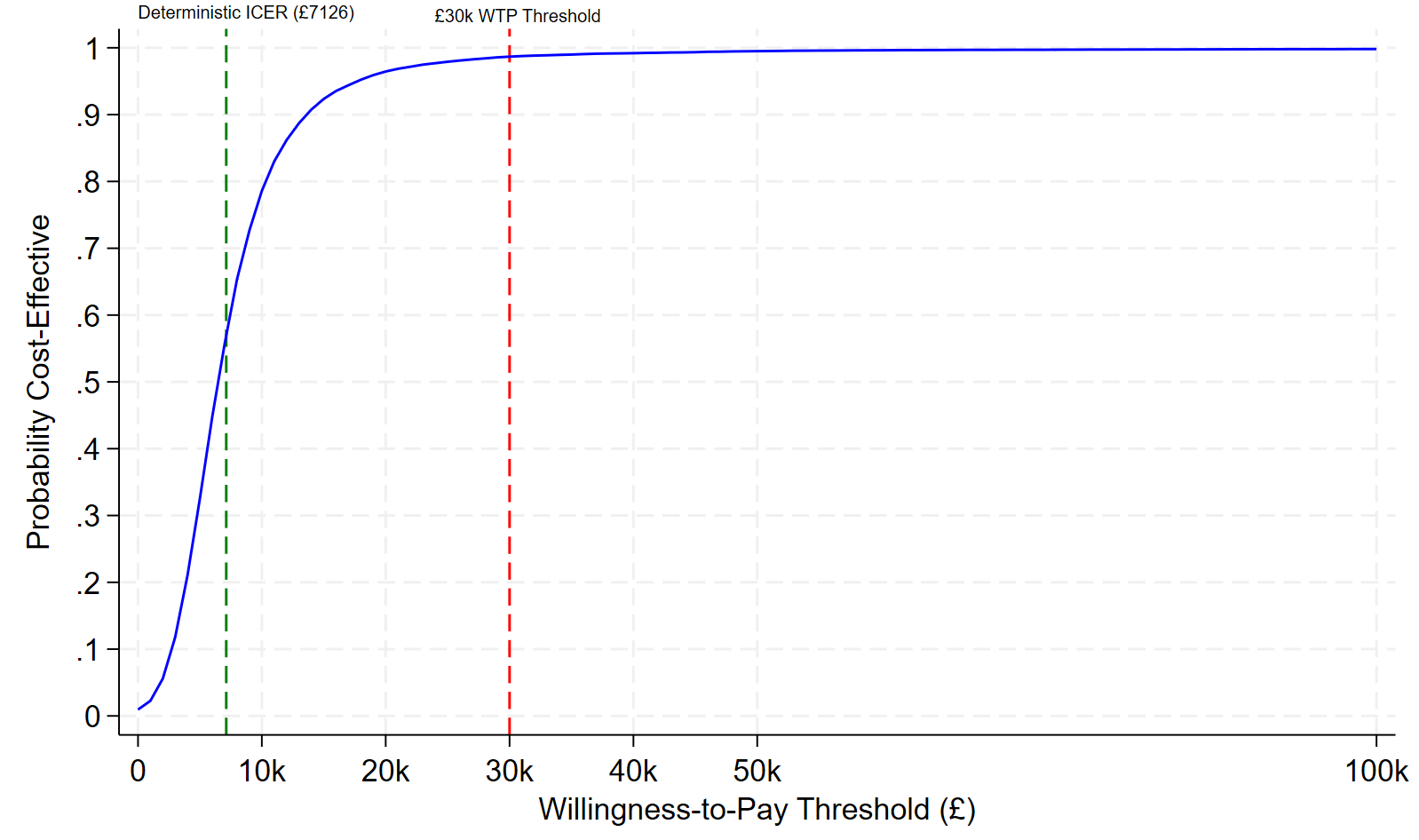

Abbreviations: ICER, incremental cost-effectiveness ratio; WTP; willingness-to-pay.

#### S17. Tornado plots showing the influence of key parameters on primary outcomes (deterministic sensitivity analysis).

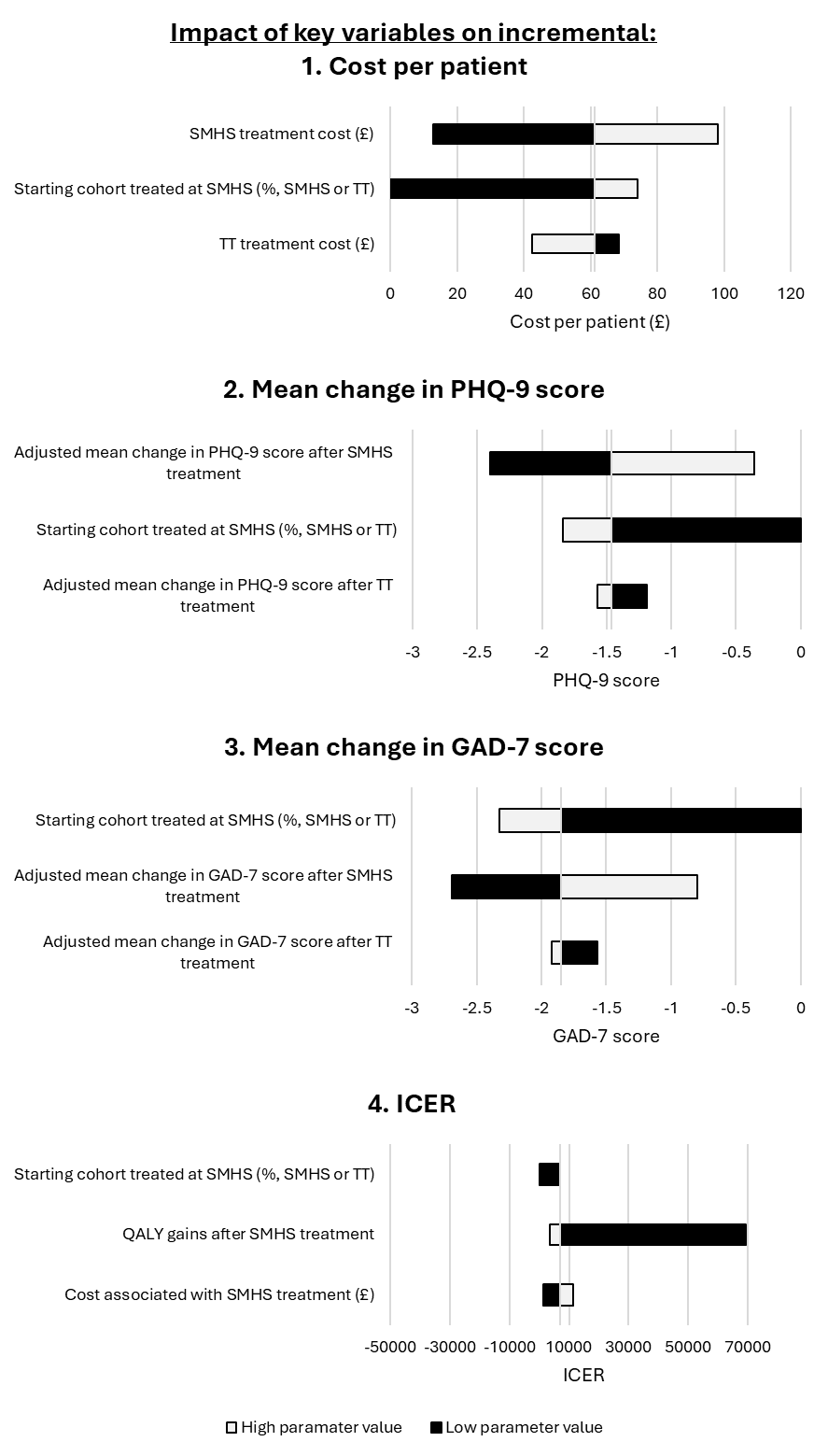

Abbreviations: GAD, Generalised Anxiety Disorder; PHQ, Patient Health Questionnaire; ICER, incremental cost-effectiveness ratio; SMHS, Staff Mental Health Service; TT, Talking Therapies.

#### S18. Sociodemographic characteristics for patient samples used in Scenario 1 – routinely collected service level data only.

|  | **Level/statistic** | **Clinical effectiveness data** | | **Resource use data** | |
| --- | --- | --- | --- | --- | --- |
|  |  | **SMHS** | **Talking Therapies** | **SMHS** | **Talking Therapies** |
| **N** |  | 48 | 1,570 | 201 | 2,093 |
| **Sex** | Female | 83.33% | 72.29% | 84.08% | 71.62% |
|  | Male | 16.67% | 27.64% | 15.92% | 28.28% |
|  | Not known | 0.00% | 0.06% | 0.00% | 0.10% |
| **Age** | Mean  (95% CI) | 38.94 (35.57, 42.30) | 36.86 (36.34, 37.39) | 38.45 (36.86, 40.04) | 37.26 (36.80, 37.71) |
| **Ethnicity** | Asian or Asian British | 2.08% | 3.69% | 4.98% | 4.30% |
|  | Black or Black British | 4.17% | 1.02% | 2.49% | 1.10% |
|  | Mixed | 2.08% | 3.18% | 1.49% | 2.87% |
|  | Other | 16.67% | 1.15% | 16.92% | 0.81% |
|  | White - British | 43.75% | 73.82% | 47.26% | 68.71% |
|  | White - Other | 8.33% | 10.06% | 6.47% | 9.99% |
|  | Not known | 22.92% | 7.07% | 20.40% | 12.23% |
| **NHS trust** | CUH | 31.25% |  | 43.78% |  |
|  | CCS | 2.08% |  | 2.49% |  |
|  | CPFT | 43.75% |  | 27.86% |  |
|  | NWAFT | 18.75% |  | 18.91% |  |
|  | RPH | 4.17% |  | 6.97% |  |
| **Staff group** | Additional Clinical Services | 20.83% |  | 19.90% |  |
|  | Additional Professional Scientific and Technical | 4.17% |  | 6.47% |  |
|  | Admin and Clerical | 18.75% |  | 15.42% |  |
|  | Allied Health Professional | 20.83% |  | 13.43% |  |
|  | Estates and Ancillary | 0.00% |  | 2.99% |  |
|  | Healthcare Scientists | 0.00% |  | 1.49% |  |
|  | Medical and Dental | 4.17% |  | 6.47% |  |
|  | Nursing and Midwifery | 27.08% |  | 23.88% |  |
|  | Not known | 4.17% |  | 9.95% |  |

All statistics have been rounded to the nearest integer; consequently, percentages may not sum to 100%.

Grey boxes indicate data not applicable.

Sex variable is dichotomised as ‘Female’ or ‘Male’ by NHS data systems.

Abbreviations: CI, confidence interval; CUH, Cambridge University Hospital; CCS, Cambridgeshire Community Services NHS Trust; CUH, Cambridge University Hospitals NHS Foundation Trust; NWAFT, North West Anglia NHS Foundation Trust; RPH, Royal Papworth Hospital NHS Foundation Trust; SMHS, Staff Mental Health Service.

#### S19. Resource use and associated costs for patient samples used in Scenario 1 – routinely collected service level data only.

|  | **SMHS** | **Talking Therapies** |
| --- | --- | --- |
| N | 201 | 2,093 |
| Therapeutic contacts | 9.23 (7.66, 10.80) | 9.46 (9.24, 9.68) |
| Total cost (unadjusted) | £650.02 (£525.90, £774.14) | £553.78 (£540.27, £567.28) |
| Total cost (adjusted^*^) | £655.18 (£596.78, £713.59) | £553.28 (£536.05, £570.51) |

All statistics reported as means with 95% confidence intervals in brackets.

Costs reported in 2022 £ GBP values.

Abbreviations: SMHS, Staff Mental Health Service.

^*^Mean total cost adjusted for age, ethnicity, and sex.

#### S20. Clinical measurements for patient samples used in Scenario 1 – routinely collected service level data only.

|  | **SMHS** | | **Talking Therapies** |
| --- | --- | --- | --- |
| N |  | |  |
| **PHQ-9** |  |  | |
| Pre-treatment | 16.90 (15.09, 18.70) | | 12.05 (11.74, 12.36) |
| Post-treatment | 9.81 (7.94, 11.68) | | 9.38 (9.07, 9.68) |
| *Change (unadjusted)* | *-7.09 (-9.69, -4.49)* | | *-2.67 (-3.11, -2.23)* |
| *Change (adjusted^*^)* | *-4.89 (-6.43, -3.37)* | | *-2.74 (-2.99, -2.48)* |
| **GAD-7** |  |  | |
| Pre-treatment | 14.19 (12.87, 15.50) | | 11.57 (11.30, 11.85) |
| Post-treatment | 7.50 (6.03, 8.97) | | 8.89 (8.61, 9.18) |
| *Change (unadjusted)* | *-6.69 (-8.66, -4.72)* | | *-2.68 (-3.08, -2.28)* |
| *Change (adjusted^*^)* | *-5.58 (-6.99, -4.17)* | | *-2.71 (-2.95, -2.47)* |
| **QALY gains** | | | |
| *Unadjusted* | *0.46 (0.34, 0.58)* | | *0.33 (0.32, 0.34)* |
| *Adjusted^*^* | *0.34 (0.33, 0.36)* | | *0.33 (0.33, 0.33)* |

All statistics reported as mean scores with 95% confidence intervals in brackets.

Abbreviations: GAD-7, Generalised Anxiety Disorder; PHQ-9, Patient Health Questionnaire; QALY, quality-adjusted life year; SMHS, Staff Mental Health Service.

^*^Mean change in scores adjusted for pre-treatment scores (pre-treatment utility for QALY gains), age, ethnicity, and sex.

#### S21. Descriptive statistics for patient samples used in Scenario 2 – complete case analysis.

|  | **Level/statistic** | **Clinical effectiveness and  resource use data** | |
| --- | --- | --- | --- |
|  |  | **SMHS** | **Talking Therapies** |
| **N** |  | 51 | 1,554 |
| **Sex** | Female | 84.31% | 72.46% |
|  | Male | 15.69% | 27.48% |
|  | Not known | 0.00% | 0.06% |
| **Age** | Mean  (95% CI) | 38.00 (34.77, 41.23) | 36.86 (36.33, 37.39) |
| **Ethnicity** | Asian or Asian British | 1.96% | 3.67% |
|  | Black or Black British | 3.92% | 1.03% |
|  | Mixed | 1.96% | 3.22% |
|  | Other | 17.65% | 1.16% |
|  | White - British | 45.10% | 73.68% |
|  | White - Other | 5.88% | 10.17% |
|  | Not known | 23.53% | 7.08% |
| **NHS trust** | CUH | 27.45% |  |
|  | CCS | 3.92% |  |
|  | CPFT | 45.10% |  |
|  | NWAFT | 17.65% |  |
|  | RPH | 5.88% |  |
| **Staff group** | Additional Clinical Services | 15.69% |  |
|  | Additional Professional Scientific and Technical | 9.80% |  |
|  | Admin and Clerical | 19.61% |  |
|  | Allied Health Professional | 15.69% |  |
|  | Estates and Ancillary | 1.96% |  |
|  | Healthcare Scientists | 0.00% |  |
|  | Medical and Dental | 3.92% |  |
|  | Nursing and Midwifery | 27.45% |  |
|  | Not known | 5.88% |  |

All statistics have been rounded to the nearest integer; consequently, percentages may not sum to 100%.

Grey boxes indicate data not applicable.

Abbreviations: CI, confidence interval; CUH, Cambridge University Hospital; CCS, Cambridgeshire Community Services NHS Trust; CUH, Cambridge University Hospitals NHS Foundation Trust; NWAFT, North West Anglia NHS Foundation Trust; RPH, Royal Papworth Hospital NHS Foundation Trust; SMHS, Staff Mental Health Service.

#### S22. Resource use and associated costs for patient samples used in Scenario 2 – complete case analysis.

|  | **SMHS** | **Talking Therapies** |
| --- | --- | --- |
| N |  |  |
| Therapeutic contacts | 12.55 (9.04, 16.05) | 9.35 (9.10, 9.60) |
| Total cost (unadjusted) | £942.82 (£626.65, £1,258.99) | £546.86 (£531.45, £562.28) |
| Total cost (adjusted^*^) | £933.67 (£829.72, £1,037.62) | £547.16 (£528.96, £565.37) |

All statistics reported as means with 95% confidence intervals in brackets.

Costs reported in 2022 £ GBP values.

Abbreviations: SMHS, Staff Mental Health Service.

^*^Mean total cost adjusted for age, ethnicity, and sex.

#### S23. Clinical measurements for patient samples used in Scenario 2 – complete case analysis.

|  | **SMHS** | | **Talking Therapies** |
| --- | --- | --- | --- |
| N |  | |  |
| **PHQ-9** |  |  | |
| Pre-treatment | 17.31 (15.67, 18.96) | | 12.03 (11.72, 12.34) |
| Post-treatment | 9.71 (7.83, 11.59) | | 9.33 (9.03, 9.64) |
| *Change (unadjusted)* | *-7.60 (-10.10, -5.10)* | | *-2.70 (-3.14, -2.26)* |
| *Change (adjusted^*^)* | *-5.23 (-6.73, -3.73)* | | *-2.78 (-3.04, -2.52)* |
| **GAD-7** |  |  | |
| Pre-treatment | 14.67 (13.43, 15.91) | | 11.56 (11.29, 11.84) |
| Post-treatment | 7.65 (6.11, 9.19) | | 8.86 (8.58, 9.14) |
| *Change (unadjusted)* | *-7.02 (-9.00, -5.04)* | | *-2.70 (-3.09, -2.31)* |
| *Change (adjusted^*^)* | *-5.69 (-7.08, -4.31)* | | *-2.75 (-2.51, -2.99)* |
| **QALY gains** | | | |
| *Unadjusted* | *0.43 (0.33, 0.54)* | | *0.33 (0.32, 0.34)* |
| *Adjusted^*^* | *0.35 (0.34, 0.36)* | | *0.33 (0.33, 0.34)* |

All statistics reported as mean scores with 95% confidence intervals in brackets.

Abbreviations: GAD-7, Generalised Anxiety Disorder; PHQ-9, Patient Health Questionnaire; QALY, quality-adjusted life year; SMHS, Staff Mental Health Service.

^*^Mean change in scores adjusted for pre-treatment scores (pre-treatment utility for QALY gains), age, ethnicity, and sex.

#### S24. Descriptive statistics for patient samples used in Scenario 3 – ‘caseness’ thresholds for symptoms of depression and anxiety.

|  | **Level/statistic** | **Clinical effectiveness and  resource use data** | |
| --- | --- | --- | --- |
|  |  | **SMHS** | **Talking Therapies** |
| **N** |  | 43 | 896 |
| **Sex** | Female | 81.40% | 73.10% |
|  | Male | 18.60% | 26.79% |
|  | Not known | 0.00% | 0.11% |
| **Age** | Mean  (95% CI) | 37.14 (33.69, 40.59) | 36.24 (35.53, 36.95) |
| **Ethnicity** | Asian or Asian British | 2.33% | 3.46% |
|  | Black or Black British | 4.65% | 0.89% |
|  | Mixed | 0.00% | 2.79% |
|  | Other | 16.28% | 1.45% |
|  | White - British | 48.84% | 75.11% |
|  | White - Other | 4.65% | 9.71% |
|  | Not known | 23.26% | 6.58% |
| **NHS trust** | CUH | 25.58% |  |
|  | CCS | 4.65% |  |
|  | CPFT | 44.19% |  |
|  | NWAFT | 18.60% |  |
|  | RPH | 6.98% |  |
| **Staff group** | Additional Clinical Services | 13.95% |  |
|  | Additional Professional Scientific and Technical | 11.63% |  |
|  | Admin and Clerical | 23.26% |  |
|  | Allied Health Professional | 11.63% |  |
|  | Estates and Ancillary | 2.33% |  |
|  | Healthcare Scientists | 0.00% |  |
|  | Medical and Dental | 4.65% |  |
|  | Nursing and Midwifery | 27.91% |  |
|  | Not known | 4.66% |  |

All statistics have been rounded to the nearest integer; consequently, percentages may not sum to 100%.

Grey boxes indicate data not applicable.

Abbreviations: CI, confidence interval; CUH, Cambridge University Hospital; CCS, Cambridgeshire Community Services NHS Trust; CUH, Cambridge University Hospitals NHS Foundation Trust; NWAFT, North West Anglia NHS Foundation Trust; RPH, Royal Papworth Hospital NHS Foundation Trust; SMHS, Staff Mental Health Service.

#### S25. Resource use and associated costs for patient samples used in Scenario 3 – ‘caseness’ thresholds for symptoms of depression and anxiety.

|  | **SMHS** | **Talking Therapies** |
| --- | --- | --- |
| N |  |  |
| Therapeutic contacts | 11.91 (8.03, 15.79) | 9.25 (8.91, 9.59) |
| Total cost (unadjusted) | £822.20 (£553.34, £1,091.05) | £548.19 (£526.59, £569.79) |
| Total cost (adjusted^*^) | £824.92 (£709.21, £940.62) | £548.06 (£523.60, £572.51) |

All statistics reported as means with 95% confidence intervals in brackets.

Costs reported in 2022 £ GBP values.

Abbreviations: SMHS, Staff Mental Health Service.

^*^Mean total cost adjusted for age, ethnicity, and sex.

#### S26. Clinical measurements for patient samples used in Scenario 3 – ‘caseness’ thresholds for symptoms of depression and anxiety.

|  | **SMHS** | | **Talking Therapies** |
| --- | --- | --- | --- |
| N |  | |  |
| **PHQ-9** |  |  | |
| Pre-treatment | 19.05 (17.67, 20.43) | | 16.13 (15.85, 16.41) |
| Post-treatment | 10.26 (8.15, 12.36) | | 11.56 (11.16, 11.97) |
| *Change (unadjusted)* | *-8.79 (-11.33, -6.25)* | | *-4.57 (-5.06, -4.08)* |
| *Change (adjusted^*^)* | *-7.24 (-8.89, -5.60)* | | *-4.26 (-4.61, -3.91)* |
| **GAD-7** |  |  | |
| Pre-treatment | 15.65 (14.60, 16.71) | | 14.94 (14.69, 15.19) |
| Post-treatment | 8.05 (6.38, 9.71) | | 10.70 (10.32, 11.07) |
| *Change (unadjusted)* | *-7.60 (-9.58, -5.62)* | | *-4.24 (-4.69, -3.79)* |
| *Change (adjusted^*^)* | *-7.40 (-9.23, -5.57)* | | *-4.63 (-5.02, -4.25)* |
| **QALY gains** | | | |
| *Unadjusted* | *0.39 (0.28, 0.49)* | | *0.31 (0.30, 0.33)* |
| *Adjusted^*^* | *0.33 (0.32, 0.34)* | | *0.32 (0.31, 0.32)* |

All statistics reported as mean scores with 95% confidence intervals in brackets.

Abbreviations: GAD-7, Generalised Anxiety Disorder; PHQ-9, Patient Health Questionnaire; QALY, quality-adjusted life year; SMHS, Staff Mental Health Service.

^*^Mean change in scores adjusted for pre-treatment scores (pre-treatment utility for QALY gains), age, ethnicity, and sex.

#### S27. Box plots comparing waiting times between SMHS and TT.

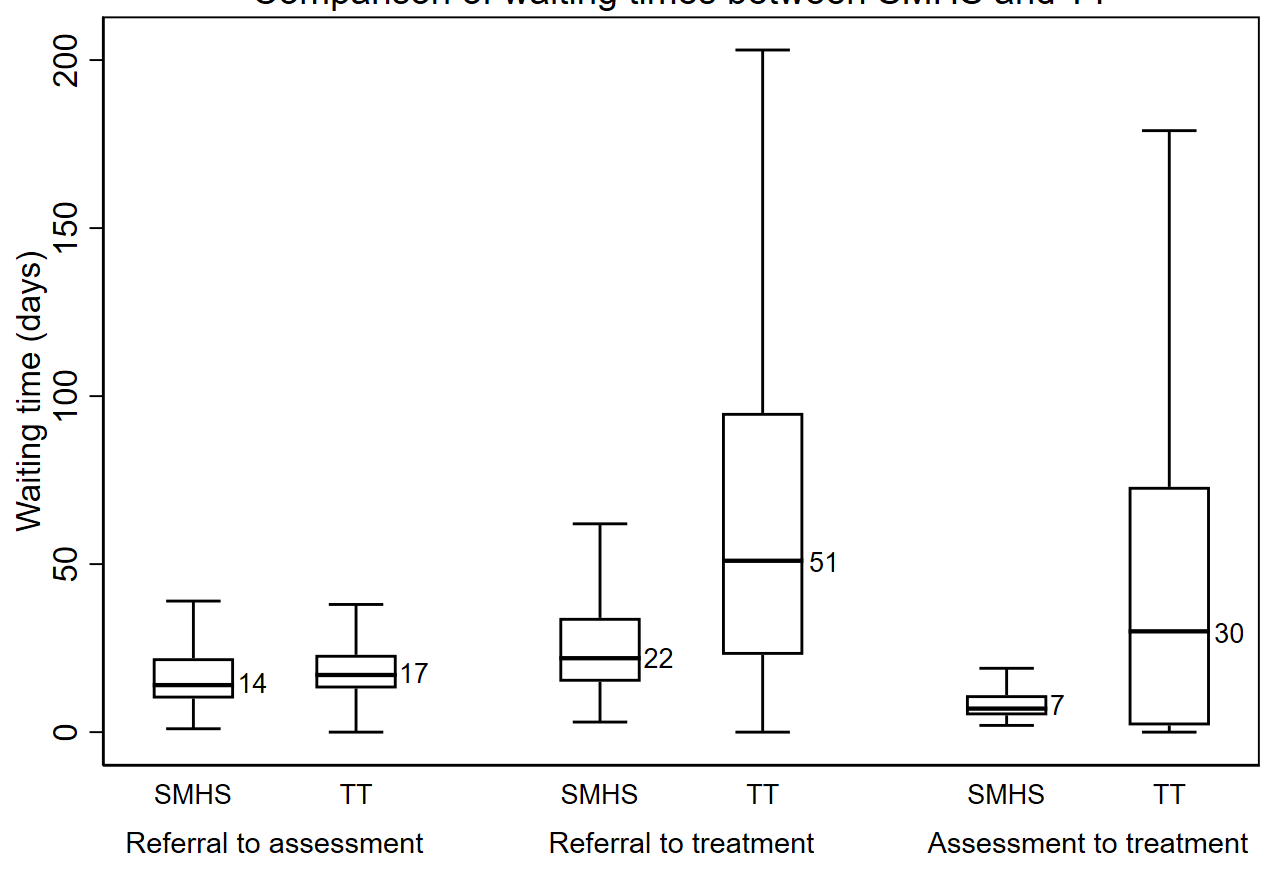

Abbreviations: SMHS, Staff Mental Health Service; TT, Talking Therapies.

Median number of days next to boxes.

#### S28. Regression model of the relationship between productivity loss* and depression severity (PHQ-9), age**, sex, and hours worked per week** at baseline.

| **Parameter** | **Coefficient** | **P-value** | **95% CI** |
| --- | --- | --- | --- |
| Constant | -0.73 |  |  |
| PHQ-9 | 0.02 | 0.001 | 0.01 – 0.04 |
| Age | 0.01 | 0.004 | 0.004 – 0.02 |
| Sex (male) | 0.01 | 0.900 | -0.21 – 0.23 |
| Work hours/ week | 0.01 | 0.080 | -0.002 – 0.02 |

Abbreviations: CI, confidence interval; OLS, ordinary least squares; PHQ, Patient Health Questionnaire.

Available sample: n=49

*In this analysis productivity loss was calculated as the proportion of usual full productivity lost to both absenteeism (time missed in the past 4 weeks days due to poor mental health) and presenteeism (percent impairment at work in the past 4 weeks due to poor mental health).

**Centred on mean values (age = 37; work hours = 35)

#### S29. Regression model of the relationship between productivity loss* and anxiety severity (GAD-7), age**, sex, and hours worked per week** at baseline.

| **Parameter** | **Coefficient** | **P-value** | **95% CI** |
| --- | --- | --- | --- |
| Constant | 0.28 |  |  |
| GAD-7 | 0.02 | 0.06 | -0.001 – 0.04 |
| Age | 0.01 | 0.02 | 0.002 – 0.02 |
| Sex (male) | -0.08 | 0.50 | -0.31 – 0.15 |
| Work hours/ week | 0.01 | 0.07 | -0.001 – 0.03 |

Abbreviations: CI, confidence interval; GAD, Generalised Anxiety Disorder; OLS, ordinary least squares.

Available sample: 49

*In this analysis productivity loss was calculated as the proportion of usual full productivity lost to both absenteeism (time missed in the past 4 weeks due to poor mental health) and presenteeism (percent impairment at work in the past 4 weeks due to poor mental health).

**Centred on mean values (age = 37; work hours = 35)

#### S30. Regression model of the relationship between productivity loss* and PTSD severity (PCL-C), age**, sex, and hours worked per week** at baseline.

| **Parameter** | **Coefficient** | **P-value** | **95% CI** |
| --- | --- | --- | --- |
| Intercept | 0.41 |  |  |
| PCL-C | 0.003 | 0.3 | -0.003 – 0.009 |
| Age | 0.01 | 0.03 | 0.001 – 0.019 |
| Sex (male) | -0.09 | 0.5 | -0.34 – 0.16 |
| Work hours/ week | 0.01 | 0.09 | -0.002 – 0.03 |

Abbreviations: CI, confidence interval; OLS, ordinary least squares; PCL-C, Post-Traumatic Stress Disorder Checklist-Civilian Version.

Available sample: 49

*In this analysis productivity loss was calculated as the proportion of usual full productivity lost to both absenteeism (time missed in the past 4 weeks due to poor mental health) and presenteeism (percent impairment at work in the past 4 weeks due to poor mental health).

**Centred on mean values (age = 37; work hours = 35)

### References

1.	Staff Mental Health Service. Cambridgeshire and Peterborough NHS Foundation Trust; 2024. <https://www.cpft.nhs.uk/smhs/>.

2.	Local Services. Cambridgeshire & Peterborough Integrated Care System; 2024. <https://www.cpics.org.uk/local-services>.

3.	Trust CaPNF. Adult & Specialist Mental Health (ASMH). Cambridgeshire and Peterborough NHS Foundation Trust; 2024. <https://www.cpft.nhs.uk/adults-specialist-mental-health-asmh-/>.

4.	Husereau D, Drummond M, Augustovski F, de Bekker-Grob E, Briggs AH, Carswell C, et al. Consolidated Health Economic Evaluation Reporting Standards 2022 (CHEERS 2022) statement: updated reporting guidance for health economic evaluations. BMC Medicine. 2022; 20(1): 23.

5.	Steen S. A cost-benefit analysis of the Improving Access to Psychological Therapies programme using its key defining outcomes. J Health Psychol. 2020; 25(13-14): 2487-98.

1. ^*^A scenario using only questionnaire data was not explored due to a small sample size (n=16).
